# Introducing the Canadian Area-Level Social Determinants of Health Indicators (CASDOHI)

**DOI:** 10.1101/2025.07.11.25331136

**Authors:** Anousheh Marouzi, Charles Plante

## Abstract

There is a growing demand for social data to support evidence-based health planning in Canada. Although Statistics Canada has made small-area Canadian census data available to researchers online for over a decade, it remains underutilized due to its cumbersome data structure and the challenges in processing and linking it with other data sources. This paper introduces the Canadian Area-Level Social Determinants of Health Indicators (CASDOHI), a user-friendly dataset that includes over 100 standardized indicators at the Dissemination Area (DA) level, harmonized between the 2011, 2016, and 2021 Census Profiles. The indicators included in CASDOHI cover various factors such as income, education, labour force, housing, and ethnocultural characteristics. The dataset also features intercensal estimates, which help researchers track changes over time. CASDOHI allows researchers to examine specific social determinants of health (SDOH) and link them to health and administrative datasets without needing advanced coding or geospatial skills. This paper also provides practical guidance for integrating these indicators with health data to support health inequality research. CASDOHI addresses a gap in Canadian health research by providing an accessible and long-running dataset of small area social determinants, facilitating the study of health inequalities, and supporting evidence-based health policymaking and planning across Canada.

**Extended Abstract:** *Introduction:* The increasing demand for high-quality social data to inform local health planning and policy-making in Canada is challenged by the technical complexity of available census data. Although Statistics Canada provides small-area level census data, their use remains limited in health research due to the resources and high data skills required to work with them.

*Objectives:* This paper presents the development and use of CASDOHI, a comprehensive dataset of over 100 indicators related to social determinants of health, harmonized across the years 2011 to 2021. It explains the construction of CASDOHI, demonstrates its relevance for researching health inequalities, and offers guidance for linking it with external health datasets. The overarching aim is to support the integration of social and health data to advance research on health disparities.

*Methods:* We developed CASDOHI using publicly available Census Profiles from 2011, 2016, and 2021 provided by Statistics Canada, as well as the NHS Profile obtained through a formal request. Additionally, we estimated CASDOHI indicators for the intercensal years from 2012 to 2020 using the linear interpolation method, while controlling for boundary changes. We selected the indicators based on a review of seven Canadian deprivation indices, with additions informed by health inequality literature and frameworks. All indicators are reported at the DA level—the smallest standard geographic unit used for census dissemination.

*Results:* CASDOHI includes indicators across multiple domains, including income, education, housing, labour force, and ethnocultural characteristics. Where available, we reported the variables disaggregated by sex. This paper outlines how CASDOHI can be linked to external datasets using geographic correspondence tools such as PCCF+ and areal interpolation.

*Conclusion:* CASDOHI addresses longstanding challenges in using census data for health research by offering a preprocessed, comprehensive, and accessible dataset covering the years 2011 to 2021. It supports nuanced analyses of health inequalities and fosters evidence-informed policy-making at local, regional, and national levels.

**Highlights:**

- CASDOHI is a user-friendly, open-access dataset that provides over 100 standardized indicators of social determinants of health at the DA level, such as income, education, labour force, housing, and ethnocultural characteristics.
- We built CASDOHI using the Census Profiles 2011, 2016, 2021, and NHS Profile. By eliminating the need for manual data manipulation, we made census data more accessible, allowing users to focus on analysis and interpretation.
- We released CASDOHI in CSV files, with a separate table for each year from 2011 to 2021.
- This paper guides linking CASDOHI indicators to external datasets, such as health administrative data, to support diverse public health and research applications.

## Introduction

The role of social determinants in shaping the population’s health is increasingly recognized in public health planning and health inequality research.^1–5^ However, in Canada, the integration of social data into health planning and policymaking remains limited by persistent barriers, including a lack of readily accessible linked social and health data, as well as the need for highly specialized data processing skills. Although Statistics Canada has provided access to small-area level census data online for over a decade through its Census Profiles,^6^ this data is not easily usable. The files shared by Statistics Canada are very large and complex and are structured in a manner that frequently necessitates considerable study and data restructuring before it can be effectively combined with health data.

This paper introduces the Canadian Area-Level Social Determinants of Health Indicators (CASDOHI), a user-friendly dataset that includes over 100 indicators reported at the Dissemination Area (DA) level for the years 2011 to 2021 and derived from the 2011, 2016, and 2021 Census Profiles. Its indicators cover income, education, labour force, housing, and ethnocultural characteristics. CASDOHI aims to address the structural, computational, and financial challenges of working directly with Census Profile data by providing ready-made standardized, longitudinal indicators across multiple domains. This is in contrast to leading indicators commonly used in health research, like the Material and Social Deprivation Index of the Institut national de santé publique du Québec (INSPQ) (also popularly known as “the Pampalon Index”),^7^ which, although created from many of the indicators included in CASDOHI, only provides a combined index.

By building upon and expanding existing deprivation indices, such as the Canadian Index of Multiple Deprivation (CIMD)^8^ and the Material and Social Deprivation Index,^7^ the CASDOHI offers a more detailed and flexible approach. It provides ready-to-use indicators that enable researchers to track changes over time, focus on specific social determinants of health (SDOH), and connect data to health and administrative sources without requiring advanced coding or geospatial analysis skills. In this way, CASDOHI enhances the accessibility of social data for health inequality research and evidence-based policymaking across Canada.

## Background

### Barriers to linking social and health data in Canada

In Canada, social data can be linked to health data using two main approaches: probabilistic linkage at the individual level and geographic linkage. Probabilistic linkage connects individual records across datasets using identifiers such as name, date of birth, and postal code, allowing social and health data to be merged at the person level. Statistics Canada’s Social Data Linkage Environment (SDLE)^9^ addresses many of the technical and governance challenges involved in this process by enabling secure, privacy-protective access to linked data through the Research Data Centre (RDC) program. However, despite this infrastructure, probabilistically linked data remain underutilized, largely because they are difficult to work with. Effective use requires advanced knowledge of multiple large line-level datasets and the technical skills to clean, merge, and analyze them. In contrast, geographic linkage—matching area-level social data to individual health records based on residential postal codes—is more commonly used in health system settings. It is simpler, more cost-effective, and avoids the complex, multi-organizational agreements required to handle sensitive individual-level data. While initiatives like the Health Data Research Network (HDRN)^10^ are working to reduce these barriers, building a fully integrated pan-Canadian health and social data system remains a long-term goal.

### The use and limitations of deprivation indices

Area-based deprivation indices have been developed as more accessible alternatives for merging social and health data geographically. Notable examples include the CIMD,^8^ the Canadian Marginalization Index (Can-Marg),^11^ the Pampalon Index,^7^ the Ontario Marginalization Index (On-Marg),^12^ and the Vancouver Area Neighbourhood Deprivation Index (VANDIX).^13^ While these indices are valuable, they simplify complex social realities into single composite scores. This simplification can obscure the impact of specific factors, such as education or housing quality, and limit their usefulness in detailed health inequality research.^14–16^ Moreover, they are relatively inflexible, preventing analysts from even considering these possibilities.

### Challenges of working with Census Profile data

The long-form questionnaire of Canada’s Census of Population serves as the primary source of social and demographic data, collecting detailed information from approximately 25% of households every five years.^17^ This data is publicly accessible through Census Profiles, which provide aggregated statistics at various geographic levels.^6^ However, the complex structure of the Census Profile data can make it challenging to work with, especially for students and researchers with limited data skills. The Census Profile does not follow best practice tidy data structure standards^18^ and adheres to a simplified version of the Statistical Data and Metadata eXchange (SDMX) standard.^19^ Each observation is organized by dimensions such as geography, characteristics, and measures. While this format enhances machine readability, it enforces a univariate data structure, meaning each row represents a single characteristic for a specific geography. This arrangement makes cross-tabulation or multivariate analysis difficult without significant pre-processing, often requiring hours of manual restructuring to create usable datasets.^19^

Moreover, access to certain datasets, like the 2011 National Household Survey (NHS) Profile file at the DA level, presents another challenge. These datasets are not available for public download and must be purchased.^20^ These limitations contribute to a significant gap between the potential and actual utilization of census data for researching social determinants of health in Canada.

### Addressing the gap: CASDOHI

We developed CASDOHI to address the challenges of accessing small-area social data from the census. CASDOHI is a user-friendly, open-access dataset that provides standardized indicators of the social determinants of health (SDOH) derived from Canadian census data. By offering preprocessed indicators in a clear and consistent format, CASDOHI removes the need for manual data cleaning or manipulation, allowing users to focus on analysis and interpretation. Unlike tools such as *cancensus* or *CensusMapper*,^21^ we chose not to release CASDOHI as an R package. Instead, we prioritized delivering ready-made tables and variables that can be immediately used with any software platform. In addition, CASDOHI includes imputed indicator values for intercensal years—something not typically available in existing R packages. We also focused on constructing variable series that are central to health inequalities research and ensured that these are harmonized over time. This involved controlling for boundary changes between censuses for different variables, including counts, percentages, means, and medians, a technically demanding and labour-intensive step (see Appendix A).^22^ Although CASDOHI is not released as an R package, the tables were built in R, and we provide the full code so others can reproduce, refine, or build upon our work.

In this paper, we provide a comprehensive guide on how to use CASDOHI in various situations and with different types of data. Our aim in developing CASDOHI is to enhance access to localized social data and facilitate its integration into health research and planning.

### <u>Methods</u> Data sources

We constructed the CASDOHI using the Census Profiles from Statistics Canada for the years 2011, 2016, and 2021.^6,23,24^ The Census Profiles from 2016 and 2021 offer publicly available aggregate data at various geographic levels, based on both short-form and long-form census responses. In 2011, the long-form Census was replaced by the voluntary National Household Survey (NHS). Consequently, the 2011 Census Profile contains information solely from the short-form questionnaires, while the NHS Profile is provided in separate files.^24,25^ It is worth noting that NHS data is not available for public download at the DA level.^25^ To address this limitation and ensure a comprehensive dataset, we combined the 2011 Census Profile with the DA-level NHS Profile, which we obtained directly from Statistics Canada through its Electronic File Transfer System. However, the NHS Profile we received had a different structure compared to the other Census Profiles. To maintain consistency across all years and facilitate accurate comparisons, we harmonized the 2011 data with the corresponding data from 2016 and 2021.

### Indicator selection

Each Census Profile contains over 2,000 fields that cover various socioeconomic and ethnocultural factors. To make this data more accessible for research purposes, we selected and developed over 50 indicators for inclusion in CASDOHI. Two primary criteria guided our selection. First, we included indicators documented in existing Canadian deprivation indices. To achieve this, we reviewed seven established Canadian deprivation indices: the CIMD at national, provincial, and regional levels;^8^ the Can-Marg;^11^ the Pampalon Index;^7,26^ the Manitoba Centre for Health Policy (MCHP) Social and Material Deprivation Indices;^27^ the On-Marg;^12^ the VANDIX;^13^ and the Canadian Social Environment Typology (CanSET).^28^ The indices are created using a range of indicators sourced from Canadian Census data, which demonstrate well-established connections between socioeconomic conditions and health outcomes. For a comprehensive list of the deprivation indices reviewed and the indicators used to create them, please refer to Appendix B.

Additionally, we included several variables identified in the broader health inequality literature and social determinants of health frameworks.^29,30^ This approach ensures that CASDOHI aligns with indicators that are already validated, widely used, and relevant to policy making.

### Indicator construction

#### Calculating indicators for census years

For the years 2011, 2016, and 2021, we derived the CASDOHI indicators directly from the corresponding Census Profiles. In constructing each indicator, we adhered to the measurement approaches used in previous studies and deprivation index developments to ensure methodological consistency and comparability across different studies and datasets. Depending on the type of indicator, we utilized various underlying population groups to align with practices found in the reviewed indices and health inequality literature. These reference groups included the total population, the total population excluding institutionalized residents, individuals living in private households, total private households, and individuals aged 15 years and older.

Appendix C outlines the variables available in CASDOHI, detailing each indicator’s corresponding Census Profile fields, the relevant reference populations, and the Profile ID^1^ combinations from the 2011, 2016, and 2021 datasets used for their derivation.

#### Estimating indicators for intercensal years

The Canadian Census is conducted every five years, which creates temporal gaps in the availability of annual data required for monitoring social indicators. To estimate CASDOHI for the intercensal years, we used a linear interpolation method to generate the necessary Census Profile data. This approach allows for the estimation of indicators for years not directly covered by a census, assuming a uniform rate of change between two known census years.

To implement this method, we first harmonized the census geographic units across different census years, addressing boundary changes and data continuity across census cycles. This step was necessary to ensure the comparability of DAs over time. Details on how we managed boundary changes using Statistics Canada’s Correspondence Files are provided in Appendix A.^31,32^

After harmonizing geographies, we interpolated Census Profile variables for the intercensal years using data from the two adjacent census cycles. We applied this approach to each variable in the Census Profile, which could represent a count, percentage, mean, or median that is used in developing the CASDOHI. For example, to interpolate between the 2011 and 2016 censuses, we estimated the variable for intercensal periods *x_t_* where *t* ∈ {2012, 2013, 2014, 2015} as:

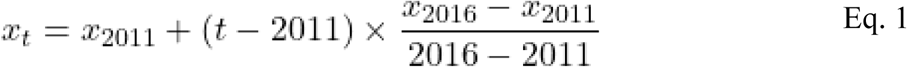

Where *x_t_* is the interpolated measure for year *t*, which could be a count, percentage, mean, or median. The values *x_2011_* and *x_2016_* are the known values from the 2011 and 2016 census years, respectively, and the denominator represents the time interval between census years.

This interpolation technique has been previously used to estimate demographic and socioeconomic data in the absence of annual census information.^22,33,34^ It provides a straightforward and transparent approach to generating annual estimates required for the development of CASDOHI for non-census years.

### Geographical unit

The CASDOHI indicators are calculated at the DA level, which is the smallest standard geographic unit used in the Canadian Census.^35^ DAs typically contain between 400 and 700 residents who share relatively similar socioeconomic characteristics, making them ideal for local policy applications.^26,35^ Additionally, the boundaries of DAs align with other census geographies, allowing for aggregation into larger units such as Census Tracts (CT), Aggregate Dissemination Areas (ADA), and Census Metropolitan Areas (CMA).^36^ Another reason for creating CASDOHI at the DA level is that most administrative health data include patients’ postal codes, which can be matched to DAs using the Postal Code Conversion File Plus (PCCF+).^37^

### Data quality and suppression

To protect privacy, Statistics Canada applies random rounding to all census counts, rounding values to the nearest 5 or 10.^38–40^ This rounding introduces minimal distortion and does not significantly impact data quality.^38^ Area suppression is also implemented under the confidentiality provisions of the Statistics Act, primarily affecting DAs with fewer than 40 residents.^38–40^ Approximately 2.5% of DAs in the Census Profiles are subject to this suppression. In CASDOHI, we considered all suppressed values as missing.

### Overview of CASDOHI indicators

The sections below are organized according to the main SDOH categories in the CASDOHI structure. Each section details the specific indicators for that category, explains the rationale behind their inclusion, and describes how they were constructed. We specifically highlight how the indicators in CASDOHI align with the variables used to develop deprivation indices and present relevant evidence from health inequalities research. It is important to note that CASDOHI also provides the indicators separately for males and females to support gender-based analyses whenever possible. An overview of these indicators is provided in Table 1.

**Table 1.**
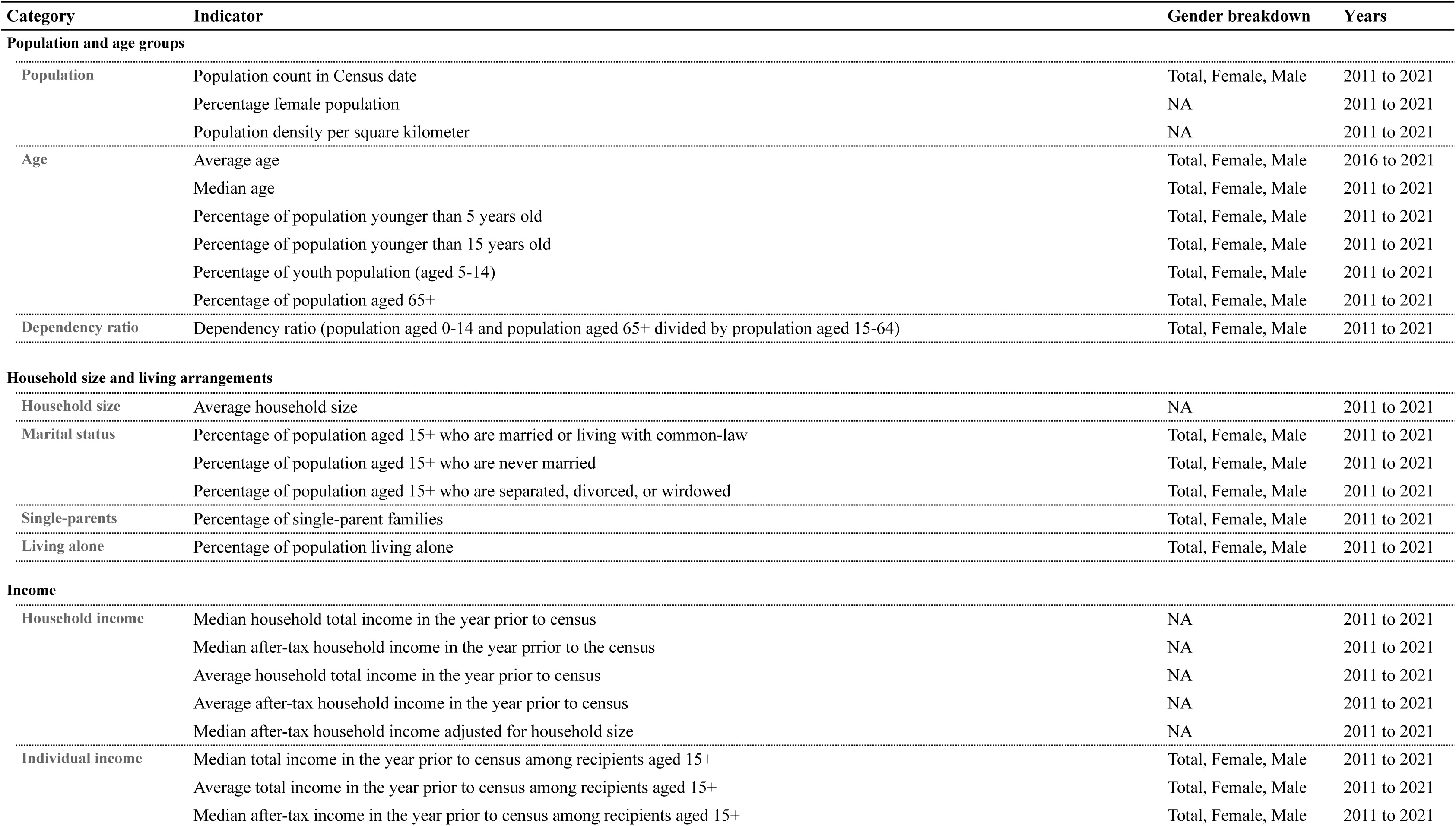
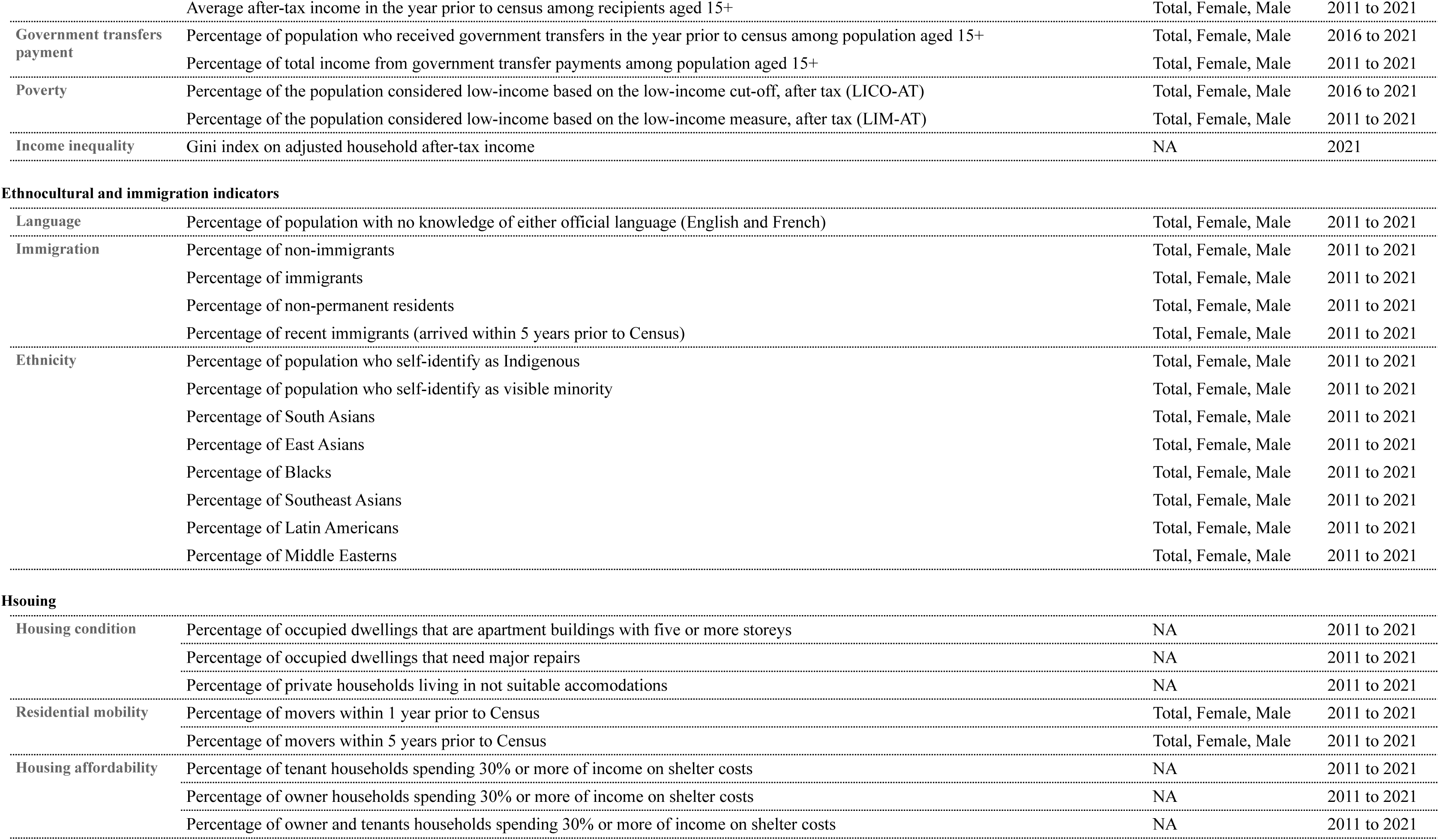
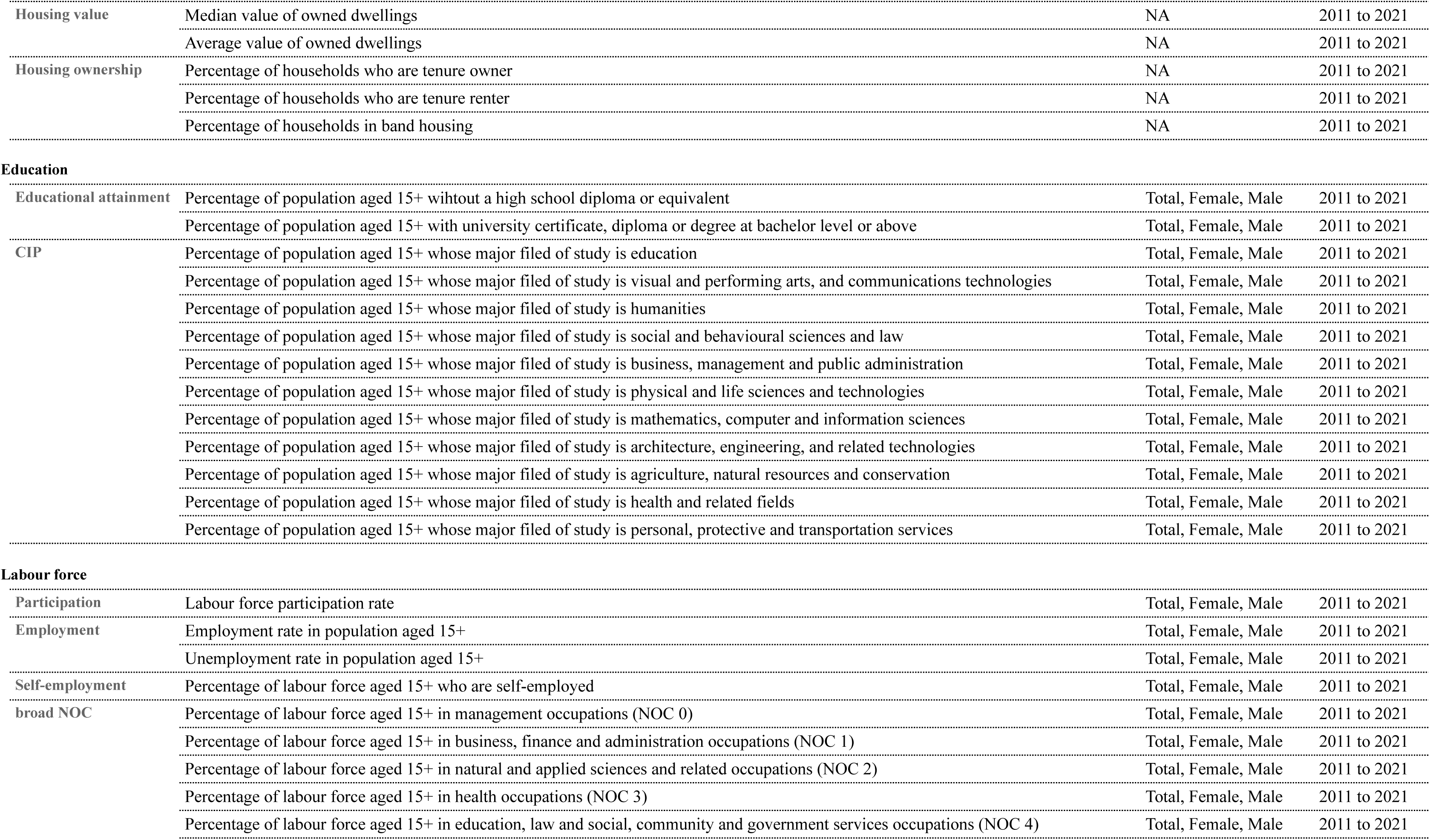
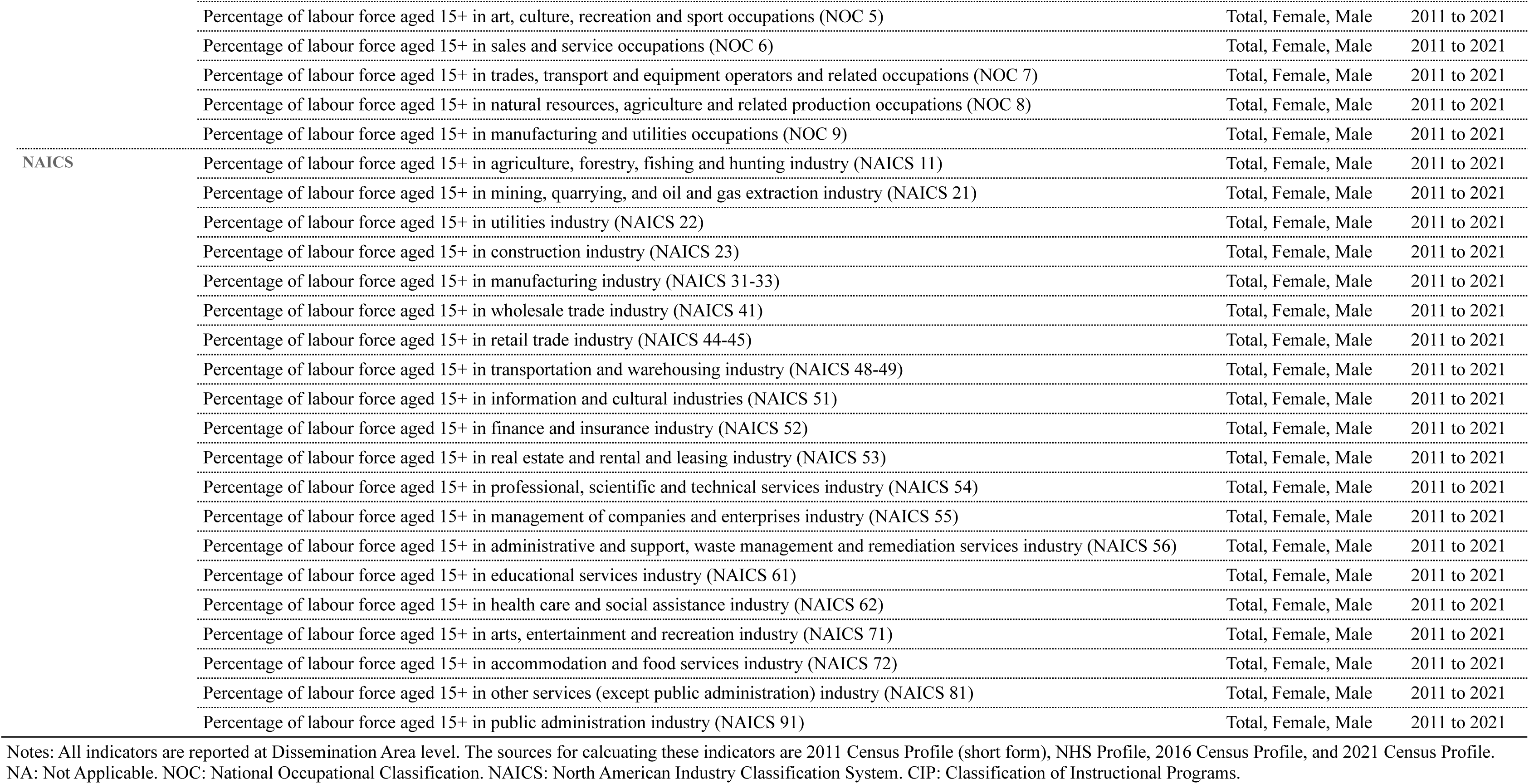
The list of indicators available in the Canadian Area-Level Social Determinants of Health Indicators (CASDOHI) dataset.

### Sex and gender

CASDOHI offers nearly all indicators broken down by total population, females, and males, enabling researchers to select the indicators that best support their specific research questions. Commonly used gender-specific indicators in CASDOHI include female unemployment, educational achievement, income, and the prevalence of female-led single-parent families.^41^ Additionally, previous studies have examined occupational characteristics by gender, which can be analyzed using CASDOHI’s occupation indicators.^41–43^ These indicators report the percentage of the population employed in various categories specified by the National Occupational Classification (NOC)^44^ and the North American Industry Classification System (NAICS)^45^ for both females and males.

Due to limitations in Census data collection, gender information in CASDOHI is reported as binary. Prior to 2021, the Census collected only male and female sex data.^46^ In the 2021 Census, a gender question was introduced; however, data at the DA level continue to be reported using binary categories (Men+ and Women+) to protect individual confidentiality.^47^

### Population and age groups

Beyond population counts, we provide population density measurements per square kilometre for each DA, which have been used in constructing the CIMD.^8^ These variables are reported as they appear in the Census Profiles.

As standard indicators in epidemiological studies, we report the median and average ages per DA, as obtained from the Census Profiles. CASDOHI also provides the population proportion by broad age groups, aligning with CIMD categories and other relevant indices.^8,11,12,28^ These include the proportion of the population under the age of 5, the proportion aged 0 to 14 years, the proportion of youth aged 5 to 14, and the proportion aged 65 and older. We also calculate the dependency ratio by dividing the combined population of individuals aged 0 to 14 and those aged 65 or older by the population aged 15 to 64. The dependency ratio was used in building CIMD,^8^ Can-Marg,^11^ and On-Marg,^12^ and reflects the reliance on the workforce within a given area.^8^

### Household composition and living arrangements

Indicators in this category provide valuable insights into household composition and social dynamics, enhancing our understanding of living arrangements and their socioeconomic and health implications. Consistent with the reviewed indices,^8,11–13,27,28,48^ CASDOHI includes the proportion of single-parent families. This is calculated as the percentage of census families led by single parents, further broken down by female and male single-parent households. Marital status is reported for individuals aged 15 and older and is categorized as: (1) married or living with a common-law partner, (2) never married, and (3) separated, divorced, or widowed. This categorization aligns with the structures used in the reviewed deprivation indices.^8,11,12,26,27,48^

Another important indicator is the proportion of individuals living alone, which is commonly used in major Canadian deprivation indices.^8,11,12,26,27,48^ In our review, we found that this measure is calculated for two groups: individuals aged 15 and older (as in the Pampalon index)^26,48^ and the entire population (as used in the MCHP, CIMD, Can-Marg, and On-Marg indices).^8,11,12,27^ Since the latter is more frequently adopted, we derive this indicator by dividing the number of single-person households by the total population in private households.

It is essential to clarify that in Census terminology, a household refers to an individual or a group of people living together in the same dwelling, regardless of their relationship. This can include both private and collective dwellings.^49^ However, the household-related indicators in CASDOHI exclude collective dwellings, as the Census Profiles report statistics solely on private households for these variables.^50^

### Income

We included median and average income in CASDOHI for individuals and households, following their availability in the Census Profile. The household income indicators are calculated for all private households within a DA, while individual income indicators apply to the population aged 15 years and older. All reviewed deprivation indices have utilized a version of these indicators in their development.^8,11–13,26–28,48^ Additionally, we included an adjusted measure of after-tax household income by dividing the median after-tax household income by the square root of the average household size. This method, derived from the CanSET,^28^ is widely recommended for assessing economic inequalities.^51,52^ After-tax household income is generally preferred over individual income because it better reflects the shared economic resources available within a household. Adjusting for household size further enhances comparability by providing a per-person estimate of financial resources.^52–54^ Moreover, median income is used instead of average income to minimize the influence of outliers and provide a more stable summary measure at the area level.

To evaluate the prevalence of low-income status, CASDOHI reports two commonly used measures: the Low-Income Cut-Offs (LICO)^55^ and the Low-Income Measure (LIM).^56^ While LICO has been employed in various indices like Can-Marg and On-Marg, LIM is generally considered a more meaningful indicator for health inequality research.^57^ These measures are reported as available in the Census Profile. Although the Market Basket Measure (MBM) is Canada’s official poverty threshold, it is not available in the Census Profile data and, therefore, could not be included.^58^

CASDOHI also incorporates the Gini coefficient, a standard indicator of income inequality in health studies.^59^ This measure is calculated at the DA level using adjusted after-tax household income and is only available from the 2021 Census.^60^

Finally, CASDOHI includes indicators of government transfers, consistent with measures used in CIMD, CanSET, Can-Marg, and On-Marg.^8,11,12,28^ These indicators refer to all cash benefits received from federal, provincial, territorial, or municipal governments and are reported both as the proportion of individuals receiving transfers and as a share of total income.

### Ethnocultural and immigration indicators

Ethnic background and immigration status are crucial factors in health inequality research. Individuals who identify as visible minorities often encounter systemic barriers, including structural racism and anti-immigrant discrimination, which lead to negative health outcomes.^61^ Many of the deprivation indices examined in this study include the proportion of the population identifying as a visible minority as a key variable.^8,11,12,28^ CASDOHI incorporates this indicator, calculated as the percentage of individuals in private households who self-identify as visible minorities, following the definition used in the Census of Population. According to the *Employment Equity Act*, visible minorities are defined as “persons, other than Aboriginal peoples, who are non-Caucasian in race or non-white in colour.”^62^

Among the indices reviewed, only CanSET disaggregates visible minority populations into subgroups, which include: (1) Arab or West Asian, (2) Latin American, (3) Black, (4) Chinese, Filipino, Southeast Asian, Korean, or Japanese, and (5) South Asian.^28^ We adhered to the Canadian Institute for Health Information (CIHI) guidelines on race-based data collection to report the proportion of the population identifying with these categories, computed as the number of individuals reporting each group divided by the total population in private households.^63^ Definitions of these categories align with CIHI’s *Guidance on the Use of Standards for Race-Based and Indigenous Identity Data Collection and Health Reporting in Canada (2022)*.^63^

In addition to visible minority status, CASDOHI includes the percentage of the population self-identifying as Indigenous, consistent with indicators included in both the CIMD and CanSET.^8,28^ This measure underscores the importance of Indigenous identity as a determinant of health and aligns with established equity frameworks.^29,30^

Immigration status is also a key determinant of health and is commonly featured in health inequality research.^61^ CASDOHI reports the percentage of the population in private households across three standard immigration categories: non-immigrants,^2^ immigrants,^3^ and non-permanent residents^4^.^6^ Some CIMD indicators use the proportion of the foreign-born population, while CanSET includes the percentage of immigrant populations in its spatial typology.^8,28^ Additionally, CASDOHI provides the percentage of recent immigrants, defined as those who arrived in Canada within five years prior to the Census, consistent with indicators used in CIMD, Can-Marg, On-Marg, and CanSET.^8,11,12,28^ These indicators help capture population groups that may face extra barriers to integration and access to services.

Finally, CASDOHI includes a measure of linguistic isolation, calculated as the percentage of the population (excluding institutional residents) who report no knowledge of Canada’s official languages—English and French. This indicator, also found in CIMD and CanSET, is widely used to highlight potential communication barriers and their implications for access to services and social inclusion.^8,28^

### Housing

Housing significantly influences health through factors such as ownership, condition, affordability, mobility, and value. These factors are prominently featured in Canadian deprivation indices. We developed a set of housing-related indicators for the CASDOHI that align with those used in these deprivation indices.^8,11–13,27,28^ CASDOHI reports the percentage of households that own their homes, providing valuable insights into ownership patterns among private households. This metric has consistently appeared in previous indices.^8,11–13,28^ To evaluate affordability, we selected the shelter-cost-to-income ratio, which indicates the proportion of a household’s total income spent on housing costs.^64^ Specifically, we included the percentage of households that spend 30% or more of their income on shelter. This measure excludes households with zero or negative income, those in band housing, and agricultural operations managed by household members. We also present this indicator separately for renters and homeowners to offer more detailed insights into affordability. These measures adhere to the methodology established by Statistics Canada and are consistent with approaches used in CIMD and CanSET.^8,28^

Indicators of housing condition and suitability encompass the percentage of private dwellings requiring major repairs and the proportion of households living in accommodations that do not meet the National Occupancy Standard (NOS), which is a benchmark for appropriate household size.^65^ These factors reflect housing adequacy and are included in indices such as CIMD, On-Marg, and CanSET.^8,12,28^ Additionally, CASDOHI considers the proportion of occupied dwellings located in apartment buildings with five or more stories, which indicates high-density urban housing environments, as identified in CIMD, Can-Marg, and On-Marg.^8,11,12^

To assess residential mobility, CASDOHI reports the percentage of individuals who changed their residence within one year and five years prior to the Census. These indicators provide insights into population stability and neighbourhood change, aligning with metrics included in CIMD, MCHP, Can-Marg, and On-Marg.^8,11,12,27^ These indicators exhibit both internal and external movers, as well as mobility within the same city.

Finally, we included two indicators related to housing value: the average and median self-reported values of owner-occupied homes. These figures serve as proxies for local housing markets and household wealth, and they have been utilized in both CIMD and CanSET.^8,28^

### Education

The health inequality literature has consistently demonstrated a strong and persistent gradient in health by educational attainment.^29,66,67^ In the various deprivation indices examined in this study, education is typically considered by the percentage of the population without a high school diploma or an equivalent certificate. Different indices use varying age cutoffs to calculate this percentage. Some consider individuals aged 25 to 64 years,^8^ while others include those aged 20 and over.^11–13^ In CASDOHI, we calculated this percentage for the population aged 15 years and older, as this approach is the most commonly used in existing indices.^11,26–28,48^ Both VANDIX and CanSET also include this indicator in their construction.^13,28^

Additionally, CASDOHI provides the percentage of the population categorized by their major field of study, based on the Classification of Instructional Programs (CIP) used in the Census.^68^ These percentages are calculated for individuals aged 15 and over, including those without a postsecondary certificate, diploma, or degree. This methodology aligns with the reporting practices utilized by Statistics Canada and allows researchers to examine health disparities across different fields of study.^69^

### Labour force

CASDOHI provides a standard set of labour market indicators commonly used in health inequality research. These indicators include labour force participation, employment, unemployment, and self-employment rates, all calculated for individuals aged 15 and older. Their inclusion aligns with the widely recognized use of these indicators in Canadian deprivation indices.^7,8,11–13,26–28,48^

In addition to these core indicators, CASDOHI offers two sets of measures that describe the composition of the employed population by occupation and industry. Both sets adhere to Statistics Canada’s reporting conventions and are expressed as percentages of the total employed population aged 15 and older.^70,71^ For occupation-based indicators, CASDOHI reports employment rates according to broad NOC categories.^44^ These rates are calculated by dividing the number of employed individuals in a specific occupational group by the total employed population. Among the indices reviewed, CanSET is the only one that incorporates occupational data, classifying the labour force into three broad categories: (1) manufacturing, construction, and trade-related occupations; (2) management and administrative occupations; and (3) professional occupations.^28^

Industry-based indicators follow a similar structure using NAICS categories.^45^ Each measure represents the percentage of the employed population working in a specific industry sector, calculated as the number of individuals in that category divided by the total employed population aged 15 and older.

### How to use CASDOHI

CASDOHI provides over 100 SDOH indicators at the DA level from 2011 to 2021. We release CASDOHI in comma-separated values (.csv) files, with a separate table for each year. The data files are released according to DA boundaries from census years 2011, 2016, and 2021, as detailed in Table 2. For the census years, we report data according to the DA boundaries of that specific census, while the boundaries from the preceding census are used to report the intercensal years. Both the CASDOHI data files and the R scripts used to generate them are publicly available on Google Drive (https://shorturl.at/4YBDy).

**Table 2.**
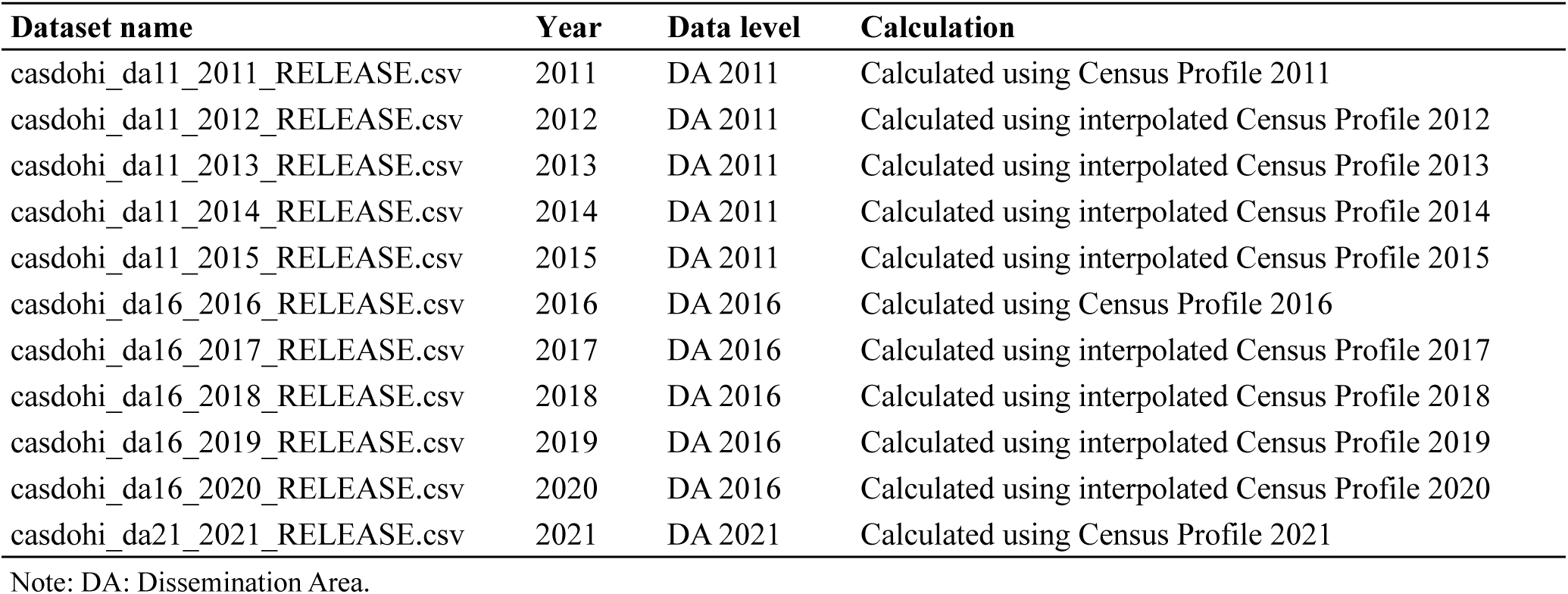
CASDOHI data products from 2011 to 2021.

| **Dataset name** | **Year** | **Data level** | **Calculation** |
| --- | --- | --- | --- |
| casdohi_da11_2011_RELEASE.csv | 2011 | DA 2011 | Calculated using Census Profile 2011 |
| casdohi_da11_2012_RELEASE.csv | 2012 | DA 2011 | Calculated using interpolated Census Profile 2012 |
| casdohi_da11_2013_RELEASE.csv | 2013 | DA 2011 | Calculated using interpolated Census Profile 2013 |
| casdohi_da11_2014_RELEASE.csv | 2014 | DA 2011 | Calculated using interpolated Census Profile 2014 |
| casdohi_da11_2015_RELEASE.csv | 2015 | DA 2011 | Calculated using interpolated Census Profile 2015 |
| casdohi_da16_2016_RELEASE.csv | 2016 | DA 2016 | Calculated using Census Profile 2016 |
| casdohi_da16_2017_RELEASE.csv | 2017 | DA 2016 | Calculated using interpolated Census Profile 2017 |
| casdohi_da16_2018_RELEASE.csv | 2018 | DA 2016 | Calculated using interpolated Census Profile 2018 |
| casdohi_da16_2019_RELEASE.csv | 2019 | DA 2016 | Calculated using interpolated Census Profile 2019 |
| casdohi_da16_2020_RELEASE.csv | 2020 | DA 2016 | Calculated using interpolated Census Profile 2020 |
| casdohi_da21_2021_RELEASE.csv | 2021 | DA 2021 | Calculated using Census Profile 2021 |

Note: DA: Dissemination Area.

This section outlines the process for linking external datasets with CASDOHI. All users should start by identifying the smallest geographic unit in their data. Depending on the type of geography, additional steps may be necessary to prepare the data for linkage. Figure 1 summarizes these steps, which are explained in detail below.

**Figure 1.**
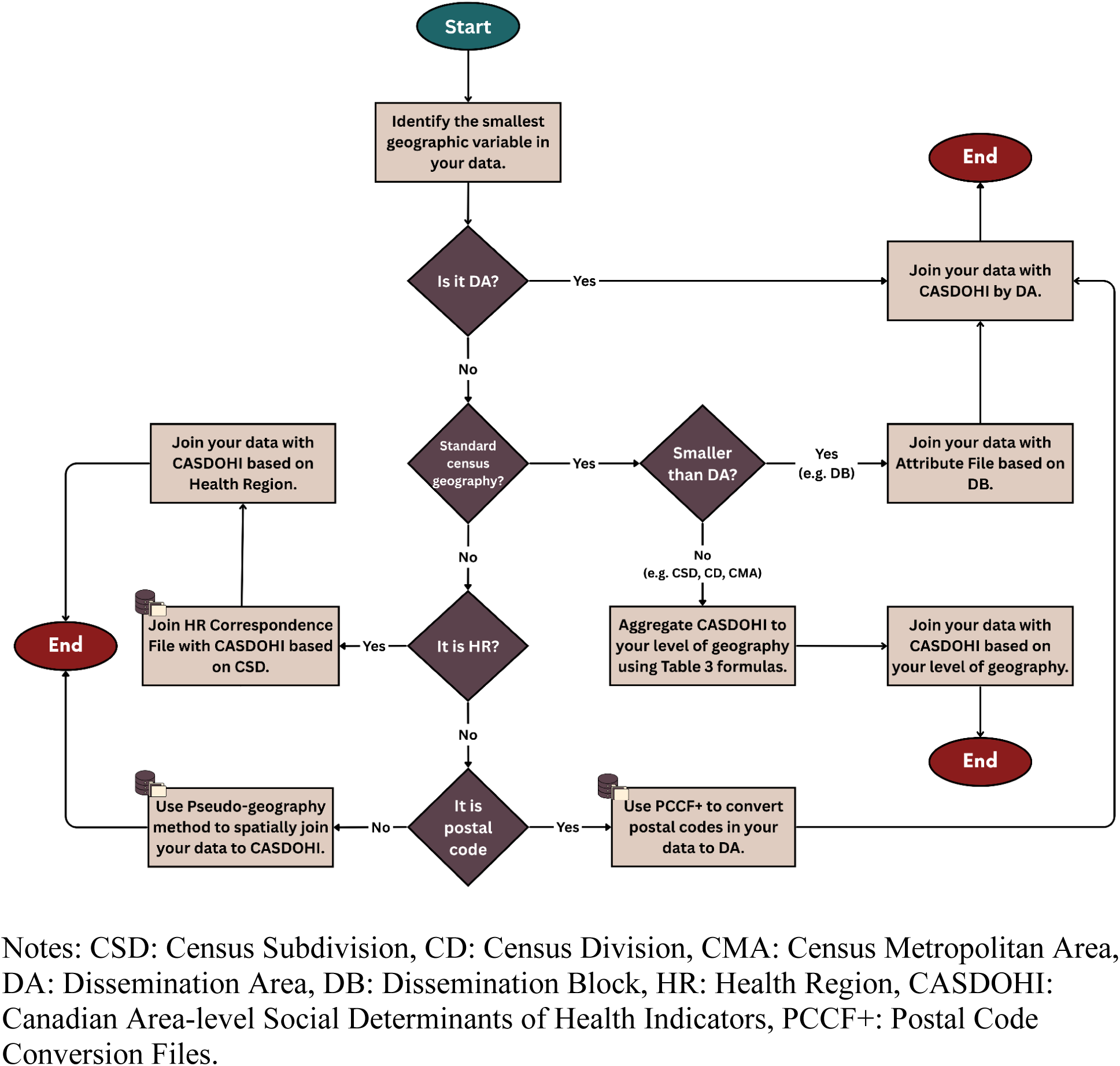
How to link CASDOHI with external datasets.

Users of CASDOHI should credit both the CASDOHI team and Statistics Canada when utilizing the dataset in publications or presentations by citing this working paper (or later iterations) and the original data sources they use.^6,23,24^

### Working with census geographies

CASDOHI provides DA-level data that can be combined with any external dataset, including geospatial information; however, how this should be done will depend on what form that geospatial information takes. If the external dataset includes a DA variable, it can be linked directly to CASDOHI without any modifications. For datasets that do not contain a DA variable, users should first determine whether it includes any other spatial unit that matches any of Statistics Canada’s standard census geographies. To do so, they can refer to Statistics Canada’s *Hierarchy of Standard Geographic Areas for Dissemination*.^36^ When working with external datasets that include Dissemination Blocks (DBs)—census units smaller than DAs—researchers should first assign DA identifiers to the DBs in their data using Statistics Canada’s Attribute File before linking it to CASDOHI. For larger census geographies, like CSD, CD, or CMA, it is essential to aggregate CASDOHI indicators to the relevant level prior to combining them with the external data.

CASDOHI encompasses six types of indicators: frequencies, percentages, ratios, means, medians, and Gini coefficients. Each of these measures requires a distinct data aggregation approach. Table 3 provides the best practices for calculating summary measures at broader geographic levels. In that, frequencies are calculated by summing values across the DAs linked to a given area. Means, percentages, and ratios require population-weighted summation. For medians, the weighted median of medians (WM) method proposed by McGrath et al.^72^ can be used, although it only approximates the population median. Due to the lack of individual-level data, the original Gini coefficient in CASDOHI cannot be precisely calculated for larger geographies. However, an extended Gini coefficient can be estimated, which accounts for both within-DA and between-DA inequalities.^73^ All necessary components for implementing these aggregation procedures are available within CASDOHI.

**Table 3.**
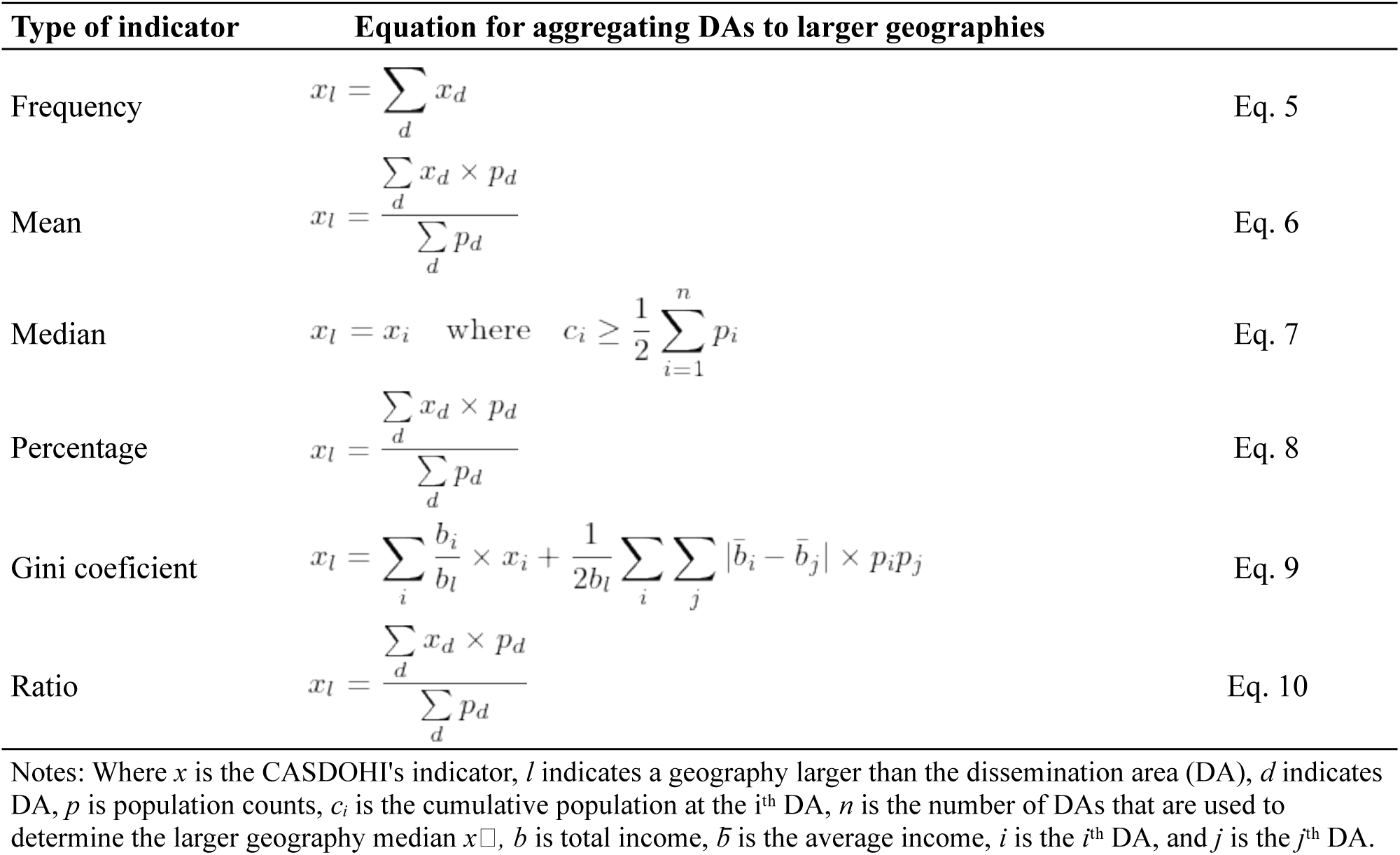
Recommended equations for aggregating DAs to larger geographies.

### Working with non-census geographies

When the smallest spatial unit in the external dataset is not one of Statistics Canada’s standard census geographies, customized tools or correspondence tables may be needed to facilitate linkage to CASDOHI. A common example in administrative health data is the use of postal codes, which can be mapped to DAs using the Postal Code Conversion File Plus (PCCF+).^37^ For datasets at the health region level, CASDOHI can first be joined to Statistics Canada’s Health Region Correspondence Files^74^ based on CSD, after which the data can be linked to the external data at the health region level. If no appropriate correspondence tool is available, we suggest using a pseudo-geography method that relies on areal interpolation.^75^ This technique allows researchers to estimate CASDOHI indicators for the geographic units they are interested in, facilitating the integration of external data at that specific level.

### Limitations

CASDOHI has several limitations that should be noted. First, not all indicators are available for every year between 2011 and 2021. For instance, the Gini coefficient is only available for 2021, and data on average age starts from 2016. These gaps arise from variations in indicator availability across Census Profile releases, as certain characteristics are not consistently included in every census cycle. Table 1 shows the availability of indicators for each year.

Second, the intercensal estimates of *population density per square kilometre* were derived using the same linear interpolation method as population counts. This method assumes an equal distribution of the population across the area, which may introduce bias. We applied this method due to the absence of land area data in Statistics Canada’s Correspondence Files.^32^ In the future, this estimation can be improved by incorporating the land area data at the DA level when performing area-weighted interpolation.

Finally, we employed areal-weighted interpolation to account for changing DA boundaries across census cycles, as this remains one of the simplest techniques available. However, more advanced spatial interpolation methods, such as population-weighted or dasymetric techniques, could enhance accuracy and should be considered in future research.^76–78^

## Conclusion

This study introduces the Canadian Area-Level Social Determinants of Health Indicators (CASDOHI), a publicly accessible dataset designed to improve access to small-area social data for health inequality research in Canada. CASDOHI addresses technical and geographic challenges associated with using census data by providing over 100 harmonized indicators at the DA level, drawn from the 2011, 2016, and 2021 Census Profiles and the 2011 NHS Profile.^6,23–25^

We selected and constructed indicators by reviewing established Canadian deprivation indices and health inequality frameworks, ensuring they align with validated measures used in prior research.^7,8,11–13,27–30,48^ CASDOHI covers key social domains—income, education, housing, labour force, and ethnocultural composition—and includes interpolated values for intercensal years to enable consistent longitudinal and area-based analyses. To enhance its practical use, we provide detailed guidance for linking CASDOHI with external datasets across various geographic formats. By lowering technical barriers, CASDOHI empowers researchers to conduct more flexible health inequality and equity-focused research.

## About the Research Department

The Saskatchewan Health Authority Research Department leads collaborative research to enhance Saskatchewan’s health and healthcare. We provide diverse research services to SHA staff, clinicians, and team members, including surveys, study design, database development, statistical analysis, and assistance with research funding. We also spearhead our own research programs to strengthen research and analytic capability and learning within Saskatchewan’s health system.

## Disclaimer

This working paper is for discussion and comment purposes. It has not been peer-reviewed nor been subject to review by Research Department staff or executives. Any opinions expressed in this paper are those of the author(s) and not those of the Saskatchewan Health Authority.

## Suggested Citation

Marouzi Anousheh, Plante Charles. 2025. “Introducing the Canadian Area-Level Social Determinants of Health Indicators (CASDOHI).” MedRxiv.

## Author Contributions

AM conducted the data analysis and prepared the first draft of the article. AM and CP designed the study and directed its implementation, including quality assurance and control. CP supervised the data analysis. CP reviewed, edited, and finalized the text. CP provided the overall guidance and funding for the research project. All authors approved the final version of the manuscript.

## Data Availability

Most of the data used in this study is publicly available and can be accessed through the Statistics Canada website. The NHS Profile at the dissemination area level is available for purchase from Statistics Canada.

https://www12.statcan.gc.ca/census-recensement/2021/dp-pd/prof/index.cfm?Lang=E

## Acknowledgments

This work builds on the exceptional efforts of Statistics Canada employees, whose dedication to collecting, processing, and disseminating Census of Population data made this study possible. We would like to express our gratitude to Samuel Price for facilitating access to the NHS Profile data, and to Dr. Jeff Allen and Dr. Zack Taylor for sharing valuable information about the Canadian Longitudinal Census Tract Database (CLTD). We also appreciate Dr. Daniel Yupanqui for his helpful comments on the formulas in this paper. Special thanks go to Dr. Suvadra Datta Gupta for reviewing the final version and providing insightful feedback. Additionally, we are grateful to Dr. Maureen Anderson and Fernando Maldonado for reviewing the manuscript and advising us on strategies for disseminating this resource within the healthcare system. The views expressed in this work are solely our own.

## Ethics Declaration

The University of Saskatchewan Research Ethics Board deemed this study exempt (# E486).

## Data Availability

Most of the data used in this study are publicly available and can be accessed through the Statistics Canada website. The NHS Profile at the dissemination area level is available for purchase from Statistics Canada.

## Code Availability

Codes are available as a supplementary file to this working paper.

## Footnotes

^1^Profile ID corresponds to the *Member ID: Profile of Dissemination Areas (2247)* variable in 2016 Census Profile and *CHARACTERISTIC_ID* in Census Profile 2021. Since there is no Profile ID variable in the 2011 datasets, we assigned these identifiers based on 2021 Census Profile.

^2^Non-immigrants are Canadian citizens by birth.

^3^Immigrants include individuals who are or have ever been landed immigrants or permanent residents, and those who acquired Canadian citizenship by naturalization.

^4^Non-immigrants are individuals with a usual place of residence in Canada but who are in the country on a work or study permit, or who have claimed refugee status.

