## Appendices A-C for "Introducing the Canadian Area-Level Social Determinants of Health Indicators (CASDOHI)"

### Appendix A. Managing boundary changes between censuses

Linear interpolation assumes that geographic boundaries remain constant over time. However, in practice, census boundaries are revised every five years to reflect demographic and administrative changes. These revisions may involve the splitting, merging, or reconfiguring of Dissemination Areas (DAs), making direct comparisons across census cycles problematic.

To address this challenge, we applied an areal-weighted interpolation method to harmonize Census Profile data between consecutive census years. Areal weighting is a common method for areal interpolation, first introduced by Markoff and Shapiro.[1,2] We employed this method by leveraging the Correspondence Files published by Statistics Canada, which provide the areal-overlap percentages between DAs from different census cycles.[3,4] For this study, we aligned the 2016 Census Profile to 2011 DA boundaries, and the 2021 Census Profile to 2016 DA boundaries. This allowed us to generate consistent Census Profiles for intercensal periods. The Census Profiles from 2012 to 2015 were estimated using DA 2011 boundaries, and those from 2017 to 2020 were estimated according to DA 2016 boundaries.

Most indicators in the Census of Population are frequencies. However, there are other types of variables in Census Profiles that we used in developing CASDOHI, including percentages, means, and medians, which must be treated differently during interpolation. Areal interpolation techniques vary depending on whether the variable is extensive or intensive.[5] Extensive variables, such as population counts, are dependent on the size of the geographic unit. In contrast, intensive variables, such as percentages and means, are not dependent on geographic size.[5] The areal-weighted interpolation method remains a widely used approach to interpolate both types of variables.[1,2]

For extensive variables such as counts, we estimated values for a target DA from an earlier census year based on overlapping DAs from the subsequent census year. For example, to estimate an extensive variable for DA in 2016 based on the boundaries of the previous census, 2011, $x_{2016 \mapsto 2011}$, we used the following formula:

$$x_{2016 \mapsto 2011} = \sum_j \left( x_{2016 \mapsto 2016} \times \frac{a_{jk}}{a_j} \right) \qquad \text{Eq. 2}$$

Where $k$ indicates a DA unit based on 2011 boundaries, $j$ represents a DA based on 2016 boundaries, $x_{2016 \mapsto 2016}$ represents the known value for DA $j$ in 2016, $a_{jk}$ is the land area of DA $j$ in 2016 that overlaps with the land area of a target DA $k$ in 2011, and $a_j$ is the total land area of DA $j$ in 2016. Since the Correspondence File does not contain information on DA areas in square kilometres, we used the same formula to interpolate *population density per square kilometre* over the intercensal years.

To estimate an intensive variable $x$ in the Census Profile, including means and percentages, in 2016 based on DA boundaries from 2011, the following equation was used:

$$x_{2016\mapsto 2011} = \frac{\sum_j (p_{2016\mapsto 2016} \times x_{2016\mapsto 2016} \times \frac{a_{jk}}{a_j})}{\sum_j (p_{2016\mapsto 2016} \times \frac{a_{jk}}{a_j})} \quad \text{Eq. 3}$$

Where $x_{2016\mapsto 2011}$ is the interpolated value for DA $k$ in 2016, $x_{2016\mapsto 2016}$ is the known mean or percentage for DA $j$ in 2016, and $p_{2016\mapsto 2016}$ indicates the population in DA $j$ in 2016.

To areally interpolate medians, we incorporated the weighted median of medians approach[6] with an areal-weighted interpolation technique.[1,2,5] Taking 2016 based on the boundaries of the previous census, 2011, as our example again, first, we sorted every 2016 DA $j$ that overlaps with the 2011 DA $k$, which we wanted to estimate the median for, by their medians. Next, we calculated the cumulative population for each DA $j$. We used the following equation to estimate the median in 2016 based on DA boundaries in 2011:

$$x_{2016\mapsto 2011} = x_{2016\mapsto 2016} \quad \text{where} \quad c_j \geq \frac{1}{2} \sum_j (p_{2016\mapsto 2016} \times \frac{a_{jk}}{a_j}) \quad \text{Eq. 4}$$

Where $x_{2016\mapsto 2011}$ is the estimated median in year 2016 in DA $k$, $x_{2016\mapsto 2016}$ is the known median in DA $j$ in 2016, and $c_j$ is the cumulative population at the $j^{th}$ DA.

**Appendix B.** Indicators used in reviewed Canadian deprivation indices and their equivalents in CASDOHI.

| | Deprivation index | | | | | | | |
|---|---|---|---|---|---|---|---|---|
| **Category** | **Pampalon Index** | **CIMD** | **MCHP** | **Can-Marg** | **On-Marg** | **VANDIX** | **CanSET** | **CASDOHI** |
| **Population and age groups** | | | | | | | | |
| **Population** | | Proportion of persons per square kilometer | | | | | Population density; total number of people per square kilometre of area | Population density per square kilometer |
| | | Proportion of population that is female | | | | | | Percentage female population |
| **Age** | | Proportion of children younger than age 6 | | | | | | Percentage of population younger than 5 years old |
| | | | | | | | Percent of population aged 0 to 14 years | Percentage of population younger than 15 years old |
| | | Proportion of population that are youth (aged 5-15) | | Proportion of the population who are not youth (age 5-15) | Proportion of the population who are not youth (age 5-15) | | | Percentage of youth population (aged 5-14) |
| | | Proportion of the population who are aged 65+ | | Proportion of the population who are aged 65+ | Proportion of the population who are aged 65+ | | Percent of population aged 65 and above | Percentage of population aged 65+ |
| **Dependency ratio** | | Dependency ratio (population aged 0-14 and population aged 65+ divided by propulation aged 15-64) | | Dependency ratio (total population 0-14 and 65+/ total population 15 to 64) | Dependency ratio (total population 0-14 and 65+ / total population 15 to 64 ) | | | Dependency ratio (population aged 0-14 and population aged 65+ divided by propulation aged 15-64) |
| **Household and living arrangements** | | | | | | | | |
| **Household size** | | Average number of persons per dwelling | | Average number of persons per dwelling/ Average household size | Average number of persons per dwelling | | Average number of people in a household | Average household size |
| | | Average number of persons per room | | | | | | Not included[1] |
| **Marital status** | | Proportion of the population that is married/common-law | | | | | | Percentage of population aged 15+ who are married or living with common-law |
| | The proportion of the population aged 15 and over who are separated, divorced or widowed | Proportion of the population that is single, divorced, separated, or widowed | the proportion of the population aged 15+ who are separated, divorced, or widowed | Proportion of the population who are single/divorced/widowed | Proportion of the population who are single/divorced/widowed | | | Percentage of population aged 15+ who are separated, divorced, or wirdowed |
| **Single-parents** | The proportion of single-parent families | Proportion of single parent families | proportion of single parent households | Proportion of families who are lone parent families | Proportion of families who are lone parent families | Proportion of lone-parent families among all census families | Percent of lone parent census families | Percentage of singe-parent families |
| **Living alone** | The proportion of the population aged 15 and over living alone | Proportion of persons living alone | the proportion of the population that lives alone | Proportion of the population living alone | Proportion of the population living alone | | | Percentage of population living alone |
| **Living in institution** | | | | | | | Percent of population that are institutionalized and residing in medical or long-term care facilities or shelters | Not included |
| **Income** | | | | | | | | |
| **Household income** | | Median household income | | | | | | Median after-tax household income in the year prrior to the census |

| Category | Deprivation index | | | | | | | CASDOHI |
|---|---|---|---|---|---|---|---|---|
| | **Pampalon Index** | **CIMD** | **MCHP** | **Can-Marg** | **On-Marg** | **VANDIX** | **CanSET** | |
| | | Average household income | average household income | | | | | Average after-tax household income in the year prior to census |
| **Individual income** | | Median income of individuals | | | | | | Median total income in the year prior to census among recipients aged 15+ |
| | | | | Average after-tax income for population aged 15+ | | | | Average after-tax income in the year prior to census among recipients aged 15+ |
| | The average personal income of the population aged 15 years and over | Average income of individuals | | | | Average total income among population 15 years and over | | Average total income in the year prior to census among recipients aged 15+ |
| **Government transfers payment** | | Proportion of population receiving government transfer payments | | | | | Percent of population aged 15+ receiving specific government transfers. | Percentage of population who received government transfers in the year prior to census among population aged 15+ |
| | | | | Proportion of total income from government transfer payments for population aged 15+ | Proportion of total income from government transfer payments for population aged 15+ | | | Percentage of total income from government transfer payments among population aged 15+ |
| **Poverty** | | Proportion of the population that is low-income | | Proportion of the population considered low-income | Proportion of the population considered low-income | | | Percentage of the population considered low-income based on LICO-AT and LIM-AT |
| **Ethnocultural and immigration indicators** | | | | | | | | |
| **Language** | | Proportion of population with no knowledge of either official language (linguistic isolation) | | | | | Percent with no knowledge of either of official languages | Percentage of population with no knowledge of either official language (English and French) |
| **Immigration** | | Proportion of population that is foreign born | | | | | Percent of immigrant population | Percentage of immigrants |
| | | Proportion of the population which are recent immigrants (arrived in five years prior to Census) | | Proportion of the population who are recent immigrants (arrived in past 5 years) | Proportion of the population who are recent immigrants (arrived in the past 5 years) | | Percent of recent immigrant population (landed in Canada between 2011 and 2016) | Percentage of recent immigrants (arrived within 5 years prior to Census) |
| **Ethnicity** | | Proportion of the population self-identified as visible minority | | Proportion of the population who self-identify as a visible minority | Proportion of the population who self- identify as a visible minority | | Percent of people who identified themselves as belonging to a visible minority group | Percentage of population who self-identify as visible minority |
| | | | | | | | Percent of people belonging to Arab or West Asian visible minority groups | Reported based on CIHI categories[2] |
| | | | | | | | Percent of people belonging to Latin American visible minority group | Reported based on CIHI categories[2] |
| | | | | | | | Percent of people belonging to Black visible minority group | Reported based on CIHI categories[2] |
| | | | | | | | Percent of people belonging to Chinese, Filipino, Southeast Asian, Korean or Japanese visible minority groups | Reported based on CIHI categories[2] |

| Category | Deprivation index | | | | | | | |
|---|---|---|---|---|---|---|---|---|
| | Pampalon Index | CIMD | MCHP | Can-Marg | On-Marg | VANDIX | CanSET | CASDOHI |
| | | | | | | | Percent of people belonging to South Asian visible minority group | Reported based on CIHI categories² |
| | | Proportion of the population identified as Indigenous | | | | | Percent of people who identified themselves with Aboriginal peoples of Canada | Percentage of population who self-identify as Indigenous |
| Religion | | Proportion of population with no religious affiliation | | | | | | Not included |
| **Housing** | | | | | | | | |
| Housing condition | | Proportion of homes needing major repairs | | | Proportion of households living in dwellings that are in need of major repair | | Percent of occupied private dwellings in need of major repairs | Percentage of occupied dwellings that need major repairs |
| | | | | | | | Percent of private households living in not suitable accommodations (according to the National Occupancy Standard) | Percentage of private households living in not suitable accomodations |
| | | Proportion of dwellings that are apartment buildings | | Proportion of dwellings that are apartments in a building with 5 or more stories | Proportion of dwellings that are apartment buildings | | | Percentage of occupied dwellings that are apartment buildings with five or more storeys |
| Housing ownership | | Proportion of dwellings that are owned | | Proportion of dwellings that are not owned | Proportion of dwellings that are not owned | Proportion of persons owing their home | Percent of private dwellings occupied by owner (Includes households that own or rent their private dwelling. Excluded is shelter occupancy on Indian reserves or setttlements.) | Percentage of households who are tenure owner |
| | | Proportion of occupied units that are rentals | | | | | | Not included |
| Housing affordability | | Proportion of tenant household spending 30% or more of household income on rent | | | | | | Percentage of tenant households spending 30% or more of income on shelter costs |
| | | | | | | | Percent of households spending more than 30% of its average total income shelter costs | Percentage of owner and tenants households spending 30% or more of income on shelter costs |
| | | Proportion of owner households spending 30% or more of household income on major payments | | | | | | Percentage of owner households spending 30% or more of income on shelter costs |
| Residential mobility | | Residential mobility (different house as 1 year ago) | | | | | | Percentage of movers within 1 year prior to Census |
| | | Proportion of movers within the past 5 years | the proportion of the population that has moved at least once in the past five years | Proportion of the population who moved during the past 5 years | Proportion of the population who moved during the past 5 years | | | Percentage of movers within 5 years prior to Census |
| Housing value | | Median dollar value of dwelling | | | | | | Median value of owned dwellings |

| Category | Pampalon Index | CIMD | MCHP | Can-Marg | On-Marg | VANDIX | CanSET | CASDOHI |
|---|---|---|---|---|---|---|---|---|
| | **Deprivation index** | | | | | | | |
| | | Average dollar value of dwelling | | | | | Average value of privately owned dwellings | Average value of owned dwellings |
| | | | | | | | Measure of variability in dwelling value within a DA | Not included |
| **Education** | | | | | | | | |
| **Educational attainment** | The proportion of the population aged 15 years and over without a high school diploma or equivalent | Proportion of the population aged 25-64 without a high-school diploma | the proportion of the population aged 15+ without high school graduation | Proportion of the population aged 15+/20+/25+ without a high-school diploma | Proportion of the population aged 20+ without a high-school diploma | Percentage of residents without high school completion in population aged 20+ | Percent of private households with the highest level of education of all its members “no certificate, degree or diploma” (aged 15+) | Percentage of population aged 15+ wihtout a high school diploma or equivalent |
| | | | | | | Percentage of residents with a university degree in population aged 20+ | Percent of private households with at least one member aged 15+ of the household having “university certificate, diploma or degree at bachelor level or above” | Percentage of population aged 15+ with university certificate, diploma or degree at bachelor level or above |
| | | Proportion of population aged 15-24 not attending school | | | | | | Not included |
| **Labour force** | | | | | | | | |
| **Participation** | | Proportion of population participating in the labour force (aged 15+) | | Proportion of the population not participating in labour force (aged 15+) | Proportion of the population not participating in labour force (aged 15+) | | | Labour force participation rate |
| **Employment** | The employment to population ratio for the population 15 years and over | Ratio of employment to population | | | | The ratio of those 15 years and over working or seeking work to the total population | | Employment rate in population aged 15+ |
| | | Proportion of population that is unemployed (aged 15+) | the unemployment rate of the population aged 15+ | Proportion of the population aged 15+ who are unemployed | Proportion of the population aged 15+ who are unemployed | Unemployment rate of population aged 15 years and over | Unemployment rate for population aged 15 years and above for population who were available for work | Unemployment rate in population aged 15+ |
| | | Unemployment rate in private households with children younger than age 6 | | | | | | Not included |
| **Self-employment** | | Proportion of population that is self-employed | | | | | | Percentage of labour force aged 15+ who are self-employed |
| **broad NOC and NAICS** | | | | | | | Percent of employed labour force in manufacturing, construction and trade related occupation (Includes population aged 15 and above who were available for work in the census reference week) | Percentage of labour force aged 15+ by NOC and NAICS |
| | | | | | | | Percent of employed labour force in management and administration occupation (Includes population aged 15 and above who were available for work in the census reference week) | Percentage of labour force aged 15 years and over by NOC and NAICS |

| Category | Deprivation index | | | | | | | |
|---|---|---|---|---|---|---|---|---|
| | Pampalon Index | CIMD | MCHP | Can-Marg | On-Marg | VANDIX | CanSET | CASDOHI |
| | | | | | | | Percent of employed labour force in professional occupation (Includes population aged 15 and above who were available for work in the census reference week) | Percentage of labour force aged 15 years and over by NOC and NAICS |
| **Unpaid work** | | Proportion of population at least 15 years old and doing unpaid housework | | | | | | Not included |
| | | Proportion of population at least 15 years old looking after children without pay | | | | | | Not included |
| | | Proportion of population at least 15 years old and providing unpaid care or assistance to seniors | | | | | | Not included |

Notes: [1] Indicates that data needed to create the indicator is not available in Census Profile. [2] *Guidance on the Use of Standards for Race-Based and Indigenous Identity Data Collection and Health Reporting in Canada* by CIHI, 2022.
CASDOHI: Canadian Area-Level Social Determinants of Health Indicators. CIMD: Canadian Indicator of Multiple Deprivation. Can-Marg: Canadian Marginalization Index. MCHP: Manitoba Centre for Health Policy Social and Material Deprivation Indices. On-Marg: Ontario Marginalization Index. VANDIX: Vancouver Area Neighbourhood Deprivation Index. CanSET: Canadian Social Environment Typology. NOC: National Occupational Classification. NAICS: North American Industry Classification System.

**Appendix C.** The list of variables available in the Canadian Area-Level Social Determinants of Health Indicators (CASDOHI) dataset.

| Variable | Description | Sample[1] | Profile Reference (2011)[2] | Profile Reference (2016)[3] | Profile Reference (2021)[4] |
|---|---|---|---|---|---|
| da_id_`year'[5] | Dissemination Area unique id in years 2011, 2016, and 2021 | NA[6] | NA | NA | NA |
| pr_id_`year' | Province unique id in years 2011, 2016, and 2021 | NA | NA | NA | NA |
| pr_name_`year' | Povince name in years 2011, 2016, and 2021 | NA | NA | NA | NA |
| csd_id_`year' | Census Subdivision unique id in years 2011, 2016, and 2021 | NA | NA | NA | NA |
| csd_name_`year' | Census Subdivision name in years 2011, 2016, and 2021 | NA | NA | NA | NA |
| cd_id_`year' | Census Division unique id in years 2011, 2016, and 2021 | NA | NA | NA | NA |
| cma_id_`year' | Census Metropolitan Area unique id in years 2011, 2016, and 2021 | NA | NA | NA | NA |
| ct_id_`year' | Census Tract unique id in years 2011, 2016, and 2021 | NA | NA | NA | NA |
| sactype_`year' | Statistical Area Classification in years 2011, 2016, and 2021 | NA | NA | NA | NA |
| pop_t | Population count in Census date - Total | 100% | 1 | 1 | 1 |
| pop_f | Population count in Census date - Female | 100% | 8 | 8 | 8 |
| pop_m | Population count in Census date - Male | 100% | 8 | 8 | 8 |
| pct_pop_f | Percentage of population who are female | 100% | 8_f/8_t×100 | 8_f/8_t×100 | 8_f/8_t×100 |
| pop_density | Population density per square kilometer | 100% | 6 | 6 | 6 |
| mean_age_t | Average age of the population - Total | 100% | Not available | 39 | 39 |
| mean_age_f | Average age of the population - Female | 100% | Not available | 39 | 39 |
| mean_age_m | Average age of the population - Male | 100% | Not available | 39 | 39 |
| med_age_t | Median age of the population - Total | 100% | 40 | 40 | 40 |
| med_age_f | Median age of the population - Female | 100% | 40 | 40 | 40 |
| med_age_m | Median age of the population - Male | 100% | 40 | 40 | 40 |
| pct_age_under5_t | Percentage of the population younger than 5 years old - Total | 100% | 10/1×100 | 10/1×100 | 10/1×100 |
| pct_age_under5_f | Percentage of the population younger than 5 years old - Female | 100% | 10/8×100 | 10/8×100 | 10/8×100 |
| pct_age_under5_m | Percentage of the population younger than 5 years old - Male | 100% | 10/8×100 | 10/8×100 | 10/8×100 |

| | | | | | |
|---|---|---|---|---|---|
| pct_age_under15_t | Percentage of the population younger than 15 years old - Total | 100% | (10+11+12)/1×100 | 35 | 35 |
| pct_age_under15_f | Percentage of the population younger than 15 years old - Female | 100% | (10+11+12)/8×100 | 35 | 35 |
| pct_age_under15_m | Percentage of the population younger than 15 years old - Male | 100% | (10+11+12)/8×100 | 35 | 35 |
| pct_age_5to14_t | Percentage of population that are youth (aged 5-14) - Total | 100% | (11+12) /1×100 | (11+12) /1×100 | (11+12) /1×100 |
| pct_age_5to14_f | Percentage of population that are youth (aged 5-14) - Female | 100% | (11+12)/8×100 | (11+12)/8×100 | (11+12)/8×100 |
| pct_age_5to14_m | Percentage of population that are youth (aged 5-14) - Male | 100% | (11+12) /8×100 | (11+12) /8×100 | (11+12) /8×100 |
| pct_age_65plus_t | Percentage of population who are aged 65+ - Total | 100% | (25+26+27+28+29)/8×100 | 37 | 37 |
| pct_age_65plus_f | Percentage of population who are aged 65+ - Female | 100% | (25+26+27+28+29)/8×100 | 37 | 37 |
| pct_age_65plus_m | Percentage of population who are aged 65+ - Male | 100% | (25+26+27+28+29)/8×100 | 37 | 37 |
| ratio_dep_t | Dependency ratio (population aged 0-14 and population aged 65+ divided by propulation aged 15-64) - Total | 100% | (10+11+12+25+26+27+28+29)/(90+91+92+93+94+95+96+97+98) | (9+24)/13 | (9+24)/13 |
| ratio_dep_f | Dependency ratio (population aged 0-14 and population aged 65+ divided by propulation aged 15-64) - Female | 100% | (10+11+12+25+26+27+28+29)/(90+91+92+93+94+95+96+97+98) | (9+24)/13 | (9+24)/13 |
| ratio_dep_m | Dependency ratio (population aged 0-14 and population aged 65+ divided by propulation aged 15-64) - Male | 100% | (10+11+12+25+26+27+28+29)/(90+91+92+93+94+95+96+97+98) | (9+24)/13 | (9+24)/13 |
| mean_hh_size | Average household size - Total | 100% | 57 | 58 | 57 |
| pct_mcl_t | Percentage of the population aged 15+ who are married or living with common-law - Total | 100% | 59/58×100 | 60/59×100 | 59/58×100 |
| pct_mcl_f | Percentage of the population aged 15+ who are married or living with common-law - Female | 100% | 59/58×100 | 60/59×100 | 59/58×100 |
| pct_mcl_m | Percentage of the population aged 15+ who are married or living with common-law - Male | 100% | 59/58×100 | 60/59×100 | 59/58×100 |
| pct_nm_t | Percentage of the population aged 15+ who are never married - Total | 100% | 67/58×100 | 64/59×100 | 67/58×100 |
| pct_nm_f | Percentage of the population aged 15+ who are never married - Female | 100% | 67/58×100 | 64/59×100 | 67/58×100 |
| pct_nm_m | Percentage of the population aged 15+ who are never married - Male | 100% | 67/58×100 | 64/59×100 | 67/58×100 |

| | | | | | |
|---|---|---|---|---|---|
| pct_sdw_t | Percentage of the population aged 15+ who are separated, divorced, or wirdowed - Total | 100% | (68+69+70)/58×100 | (65+66+67)/59×100 | (68+69+70)/58×100 |
| pct_sdw_f | Percentage of the population aged 15+ who are separated, divorced, or wirdowed - Female | 100% | (68+69+70)/58×100 | (65+66+67)/59×100 | (68+69+70)/58×100 |
| pct_sdw_m | Percentage of the population aged 15+ who are separated, divorced, or wirdowed - Male | 100% | (68+69+70)/58×100 | (65+66+67)/59×100 | (68+69+70)/58×100 |
| pct_single_parent_t | Percentage of single-parent families | 100% | 86/78×100 | 78/74×100 | 86/78×100 |
| pct_single_parent_f | Percentage of single-parent families in which the parent is woman | 100% | 87/78×100 | 79/74×100 | 87/78×100 |
| pct_single_parent_m | Percentage of single-parent families in which the parent is man | 100% | 88/78×100 | 80/74×100 | 88/78×100 |
| pct_alone_t | Percentage of population living alone - Total | 100% | 51/89×100 | 52/57×100 | 51/89×100 |
| pct_alone_f | Percentage of population living alone - Female | 100% | 51/89×100 | 52/57×100 | 51/89×100 |
| pct_alone_m | Percentage of population living alone - Male | 100% | 51/89×100 | 52/57×100 | 51/89×100 |
| pct_no_eng_fr_t | Percentage of population with no knowledge of either official language (English and French) - Total | 100% | 387/383×100 | 104/100×100 | 387/383×100 |
| pct_no_eng_fr_f | Percentage of population with no knowledge of either official language (English and French) - Female | 100% | 387/383×100 | 104/100×100 | 387/383×100 |
| pct_no_eng_fr_m | Percentage of population with no knowledge of either official language (English and French) - Male | 100% | 387/383×100 | 104/100×100 | 387/383×100 |
| med_ttinc_hh | Median household total income in the year prior to census ($) | 100% | 243 | 742 | 243 |
| med_atinc_hh | Median after-tax household income in the year prrior to the census ($) | 100% | 244 | 743 | 244 |
| mean_ttinc_hh | Average household total income in the year prior to census ($) | 100% | 252 | 751 | 252 |
| mean_atinc_hh | Average after-tax household income in the year prior to census ($) | 100% | 253 | 752 | 253 |
| med_ttinc_ind_t | Median total income in the year prior to census among recipients aged 15+ ($) - Total | 100% | 113 | 663 | 113 |
| med_ttinc_ind_f | Median total income in the year prior to census among recipients aged 15+ ($) - Female | 100% | 113 | 663 | 113 |
| med_ttinc_ind_m | Median total income in the year prior to census among recipients aged 15+ ($) - Male | 100% | 113 | 663 | 113 |
| mean_ttinc_ind_t | Average total income in the year prior to census among recipients aged 15+ ($) - Total | 25% | 128 | 674 | 128 |

| | | | | | |
|---|---|---|---|---|---|
| mean_ttinc_ind_f | Average total income in the year prior to census among recipients aged 15+ ($) - Female | 25% | 128 | 674 | 128 |
| mean_ttinc_ind_m | Average total income in the year prior to census among recipients aged 15+ ($) - Male | 25% | 128 | 674 | 128 |
| med_atinc_ind_t | Median after-tax income in the year prior to census among recipients aged 15+ ($) - Total | 25% | 115 | 665 | 115 |
| med_atinc_ind_f | Median after-tax income in the year prior to census among recipients aged 15+ ($) - Female | 25% | 115 | 665 | 115 |
| med_atinc_ind_m | Median after-tax income in the year prior to census among recipients aged 15+ ($) - Male | 25% | 115 | 665 | 115 |
| mean_atinc_ind_t | Average after-tax income in the year prior to census among recipients aged 15+ ($) - Total | 100% | 130 | 676 | 130 |
| mean_atinc_ind_f | Average after-tax income in the year prior to census among recipients aged 15+ ($) - Female | 100% | 130 | 676 | 130 |
| mean_atinc_ind_m | Average after-tax income in the year prior to census among recipients aged 15+ ($) - Male | 100% | 130 | 676 | 130 |
| med_atinc_hh_adj | Median after-tax household income adjusted for the household size | 100% | 244/sqr root(57) | 743/square root(58) | 244/sqr root(57) |
| pct_pop_gtransfer_t | Percentage of population who received government transfers in the year prior to census among population aged 15+ in private households - Total | 100% | Not available | 668/661×100 | 120/111×100 |
| pct_pop_gtransfer_f | Percentage of population who received government transfers in the year prior to census among population aged 15+ in private households - Female | 100% | Not available | 668/661×100 | 120/111×100 |
| pct_pop_gtransfer_m | Percentage of population who received government transfers in the year prior to census among population aged 15+ in private households - Male | 100% | Not available | 668/661×100 | 120/111×100 |
| pct_inc_gtransfer_t | Percentage of total income from government transfer payments among population aged 15+ in private households - Total | 100% | 151 | 690 | 151 |
| pct_inc_gtransfer_f | Percentage of total income from government transfer payments among population aged 15+ in private households - Female | 100% | 151 | 690 | 151 |
| pct_inc_gtransfer_m | Percentage of total income from government transfer payments among population aged 15+ in private households - Male | 100% | 151 | 690 | 151 |
| pct_lico_at_t | Percentage of the population considered low-income based on the low-income cut-off, after tax (LICO-AT) - Total | 100% | Not available | 867 | 360 |

| | | | | | |
|---|---|---|---|---|---|
| pct_lico_at_f | Percentage of the population considered low-income based on the low-income cut-off, after tax (LICO-AT) - Female | 100% | Not available | 867 | 360 |
| pct_lico_at_m | Percentage of the population considered low-income based on the low-income cut-off, after tax (LICO-AT) - Male | 100% | Not available | 867 | 360 |
| pct_lim_at_t | Percentage of the population considered low-income based on the low-income measure, after tax (LIM-AT) - Total | 100% | 345 | 857 | 345 |
| pct_lim_at_f | Percentage of the population considered low-income based on the low-income measure, after tax (LIM-AT) - Female | 100% | 345 | 857 | 345 |
| pct_lim_at_m | Percentage of the population considered low-income based on the low-income measure, after tax (LIM-AT) - Male | 100% | 345 | 857 | 345 |
| gini_index | Gini index on adjusted household after-tax income | 100% | Not abailable | Not available | 381 |
| pct_non_immig_t | Percentage of non-immigrants - Total | 25% | 1528/1527×100 | 1141/1140×100 | 1528/1527×100 |
| pct_non_immig_f | Percentage of non-immigrants - Female | 25% | 1528/1527×100 | 1141/1140×100 | 1528/1527×100 |
| pct_non_immig_m | Percentage of non-immigrants - Male | 25% | 1528/1527×100 | 1141/1140×100 | 1528/1527×100 |
| pct_immig_t | Percentage of immigrants - Total | 25% | 1529/1527×100 | 1142/1140×100 | 1529/1527×100 |
| pct_immig_f | Percentage of immigrants - Female | 25% | 1529/1527×100 | 1142/1140×100 | 1529/1527×100 |
| pct_immig_m | Percentage of immigrants - Male | 25% | 1529/1527×100 | 1142/1140×100 | 1529/1527×100 |
| pct_non_pr_t | Percentage of non-permanent residents - Total | 25% | 1537/1527×100 | 1150/1140×100 | 1537/1527×100 |
| pct_non_pr_f | Percentage of non-permanent residents - Female | 25% | 1537/1527×100 | 1150/1140×100 | 1537/1527×100 |
| pct_non_pr_m | Percentage of non-permanent residents - Male | 25% | 1537/1527×100 | 1150/1140×100 | 1537/1527×100 |
| pct_recent_immig_t | Percentage of recent immigrants (arrived within 5 years prior to Census) - Total | 25% | 1536/1527×100 | 1149/1140×100 | 1536/1527×100 |
| pct_recent_immig_f | Percentage of recent immigrants (arrived within 5 years prior to Census) - Female | 25% | 1536/1527×100 | 1149/1140×100 | 1536/1527×100 |
| pct_recent_immig_m | Percentage of recent immigrants (arrived within 5 years prior to Census) - Male | 25% | 1536/1527×100 | 1149/1140×100 | 1536/1527×100 |
| pct_indigenous_t | Percentage of population who self-identify as Indigenous - Total | 25% | 1403/1402×100 | 1290/1289×100 | 1403/1402×100 |
| pct_indigenous_f | Percentage of population who self-identify as Indigenous - Female | 25% | 1403/1402×100 | 1290/1289×100 | 1403/1402×100 |

| | | | | | |
|---|---|---|---|---|---|
| pct_indigenous_m | Percentage of population who self-identify as Indigenous - Male | 25% | 1403/1402×100 | 1290/1289×100 | 1403/1402×100 |
| pct_vm_t | Percentage of population who self-identify as a visible minority - Total | 25% | 1684/1683×100 | 1324/1323×100 | 1684/1683×100 |
| pct_vm_f | Percentage of population who self-identify as a visible minority - Female | 25% | 1684/1683×100 | 1324/1323×100 | 1684/1683×100 |
| pct_vm_m | Percentage of population who self-identify as a visible minority - Male | 25% | 1684/1683×100 | 1324/1323×100 | 1684/1683×100 |
| pct_south_asian_t | Percentage of South Asians - Total | 25% | 1685/1683×100 | 1325/1323×100 | 1685/1683×100 |
| pct_south_asian_f | Percentage of South Asians - Female | 25% | 1685/1683×100 | 1325/1323×100 | 1685/1683×100 |
| pct_south_asian_m | Percentage of South Asians - Male | 25% | 1685/1683×100 | 1325/1323×100 | 1685/1683×100 |
| pct_east_asian_t | Percentage of East Asians - Total | 25% | (1686+1693+1694)/1683×100 | (1326+1333+1334)/1323×100 | (1686+1693+1694)/1683×100 |
| pct_east_asian_f | Percentage of East Asians - Female | 25% | (1686+1693+1694)/1683×100 | (1326+1333+1334)/1323×100 | (1686+1693+1694)/1683×100 |
| pct_east_asian_m | Percentage of East Asians - Male | 25% | (1686+1693+1694)/1683×100 | (1326+1333+1334)/1323×100 | (1686+1693+1694)/1683×100 |
| pct_black_t | Percentage of Black - Total | 25% | 1687/1683×100 | 1327/1323×100 | 1687/1683×100 |
| pct_black_f | Percentage of Black - Female | 25% | 1687/1683×100 | 1327/1323×100 | 1687/1683×100 |
| pct_black_m | Percentage of Black - Male | 25% | 1687/1683×100 | 1327/1323×100 | 1687/1683×100 |
| pct_southeast_asian_t | Percentage of Southeast Asian - Total | 25% | (1688+1691)/1683×100 | (1328+1331)/1323×100 | (1688+1691)/1683×100 |
| pct_southeast_asian_f | Percentage of Southeast Asian - Female | 25% | (1688+1691)/1683×100 | (1328+1331)/1323×100 | (1688+1691)/1683×100 |
| pct_southeast_asian_m | Percentage of Southeast Asian - Male | 25% | (1688+1691)/1683×100 | (1328+1331)/1323×100 | (1688+1691)/1683×100 |
| pct_latin_american_t | Percentage of Latin American - Total | 25% | 1690/1683×100 | 1329/1323×100 | 1690/1683×100 |
| pct_latin_american_f | Percentage of Latin American - Female | 25% | 1690/1683×100 | 1329/1323×100 | 1690/1683×100 |
| pct_latin_american_m | Percentage of Latin American - Male | 25% | 1690/1683×100 | 1329/1323×100 | 1690/1683×100 |
| pct_middle_eastern_t | Percentage of Middle Eastern - Total | 25% | (1689+1692)/1683×100 | (1330+1332)/1323×100 | (1689+1692)/1683×100 |
| pct_middle_eastern_f | Percentage of Middle Eastern - Female | 25% | (1689+1692)/1683×100 | (1330+1332)/1323×100 | (1689+1692)/1683×100 |
| pct_middle_eastern_m | Percentage of Middle Eastern - Male | 25% | (1689+1692)/1683×100 | (1330+1332)/1323×100 | (1689+1692)/1683×100 |
| pct_apt_5plus | Percentage of occupied dwellings that are apartment buildings with five or more storeys | 100% | 47/41×100 | 43/41×100 | 47/41×100 |
| pct_major_repair | Percentage of occupied dwellings that need major repairs | 25% | 1451/1449×100 | 1653/1651×100 | 1451/1449×100 |
| pct_mover_1y_t | Percentage of movers within 1 year prior to Census - Total | 25% | 1976/1974×100 | 2232/2230×100 | 1976/1974×100 |
| pct_mover_1y_f | Percentage of movers within 1 year prior to Census - Female | 25% | 1976/1974×100 | 2232/2230×100 | 1976/1974×100 |

| | | | | | |
|---|---|---|---|---|---|
| pct_mover_1y_m | Percentage of movers within 1 year prior to Census - Male | 25% | 1976/1974×100 | 2232/2230×100 | 1976/1974×100 |
| pct_mover_5y_t | Percentage of movers within 5 years prior to Census - Total | 25% | 1985/1983×100 | 2241/2239×100 | 1985/1983×100 |
| pct_mover_5y_f | Percentage of movers within 5 years prior to Census - Female | 25% | 1985/1983×100 | 2241/2239×100 | 1985/1983×100 |
| pct_mover_5y_m | Percentage of movers within 5 years prior to Census - Male | 25% | 1985/1983×100 | 2241/2239×100 | 1985/1983×100 |
| pct_not_suitable | Percentage of private households living in not suitable accomodations | 25% | 1439/1437×100 | 1642/1640×100 | 1439/1437×100 |
| pct_shelter_cost_30plus_tenant | Percentage of tenant households spending 30% or more of income on shelter costs | 25% | 1492 | 1680 | 1492 |
| pct_shelter_cost_30plus_owner | Percentage of owner households spending 30% or more of income on shelter costs | 25% | 1484 | 1673 | 1484 |
| pct_shelter_cost_30plus_tenant_owner | Percentage of owner and tenants households spending 30% or more of income on shelter costs | 25% | 1467/1465×100 | 1669/1667×100 | 1467/1465×100 |
| med_dwelling_value | Median value of owned dwellings | 25% | 1488 | 1676 | 1488 |
| mean_dwelling_value | Average value of owned dwellings | 25% | 1489 | 1677 | 1489 |
| pct_owner | Percentage of households who are tenure owner | 25% | 1415/1414×100 | 1618/1617×100 | 1415/1414×100 |
| pct_renter | Percentage of households who are tenure renter | 25% | 1416/1414×100 | 1619/1617×100 | 1416/1414×100 |
| pct_band_housing | Percentage of households in band housing | 25% | 1417/1414×100 | 1620/1617×100 | 1417/1414×100 |
| pct_no_diploma_t | Percentage of the population aged 15+ wihtout a high school diploma or equivalent - Total | 25% | 1993/1998×100 | 1684/1683×100 | 1993/1998×100 |
| pct_no_diploma_f | Percentage of the population aged 15+ wihtout a high school diploma or equivalent - Female | 25% | 1993/1998×100 | 1684/1683×100 | 1993/1998×100 |
| pct_no_diploma_m | Percentage of the population aged 15+ wihtout a high school diploma or equivalent - Male | 25% | 1993/1998×100 | 1684/1683×100 | 1993/1998×100 |
| pct_uni_diploma_t | Percentage of the population aged 15+ with university certificate, diploma or degree at bachelor level or above - Total | 25% | 2008/1998×100 | 1692/1683×100 | 2008/1998×100 |
| pct_uni_diploma_f | Percentage of the population aged 15+ with university certificate, diploma or degree at bachelor level or above - Female | 25% | 2008/1998×100 | 1692/1683×100 | 2008/1998×100 |
| pct_uni_diploma_m | Percentage of the population aged 15+ with university certificate, diploma or degree at bachelor level or above - Male | 25% | 2008/1998×100 | 1692/1683×100 | 2008/1998×100 |

| | | | | | |
|---|---|---|---|---|---|
| pct_cip_education_t | Percentage of population aged 15+ whose major filed of study, by Classification of Instructional Programs (CIP), is Education - Total | 25% | 2032/2030×100 | 1715/1713×100 | 2032/2030×100 |
| pct_cip_education_f | Percentage of population aged 15+ whose major filed of study, by Classification of Instructional Programs (CIP), is Education - Female | 25% | 2032/2030×100 | 1715/1713×100 | 2032/2030×100 |
| pct_cip_education_m | Percentage of population aged 15+ whose major filed of study, by Classification of Instructional Programs (CIP), is Education - Male | 25% | 2032/2030×100 | 1715/1713×100 | 2032/2030×100 |
| pct_cip_art_t | Percentage of population aged 15+ whose major filed of study, by Classification of Instructional Programs (CIP), is Visual and performing arts, and communications technologies - Total | 25% | 2034/2030×100 | 1717/1713×100 | 2034/2030×100 |
| pct_cip_art_f | Percentage of population aged 15+ whose major filed of study, by Classification of Instructional Programs (CIP), is Visual and performing arts, and communications technologies - Female | 25% | 2034/2030×100 | 1717/1713×100 | 2034/2030×100 |
| pct_cip_art_m | Percentage of population aged 15+ whose major filed of study, by Classification of Instructional Programs (CIP), is Visual and performing arts, and communications technologies - Male | 25% | 2034/2030×100 | 1717/1713×100 | 2034/2030×100 |
| pct_cip_humanities_t | Percentage of population aged 15+ whose major filed of study, by Classification of Instructional Programs (CIP), is Humanities - Total | 25% | 2037/2030×100 | 1720/1713×100 | 2037/2030×100 |
| pct_cip_humanities_f | Percentage of population aged 15+ whose major filed of study, by Classification of Instructional Programs (CIP), is Humanities - Female | 25% | 2037/2030×100 | 1720/1713×100 | 2037/2030×100 |
| pct_cip_humanities_m | Percentage of population aged 15+ whose major filed of study, by Classification of Instructional Programs (CIP), is Humanities - Male | 25% | 2037/2030×100 | 1720/1713×100 | 2037/2030×100 |
| pct_cip_social_t | Percentage of population aged 15+ whose major filed of study, by Classification of Instructional Programs (CIP), is Social and behavioural sciences and law - Total | 25% | 2046/2030×100 | 1729/1713×100 | 2046/2030×100 |
| pct_cip_social_f | Percentage of population aged 15+ whose major filed of study, by Classification of Instructional Programs (CIP), is Social and behavioural sciences and law - Female | 25% | 2046/2030×100 | 1729/1713×100 | 2046/2030×100 |

| | | | | | |
|---|---|---|---|---|---|
| pct_cip_social_m | Percentage of population aged 15+ whose major filed of study, by Classification of Instructional Programs (CIP), is Social and behavioural sciences and law - Male | 25% | 2046/2030×100 | 1729/1713×100 | 2046/2030×100 |
| pct_cip_buisiness_t | Percentage of population aged 15+ whose major filed of study, by Classification of Instructional Programs (CIP), is Business, management and public administration - Total | 25% | 2054/2030×100 | 1737/1713×100 | 2054/2030×100 |
| pct_cip_buisiness_m | Percentage of population aged 15+ whose major filed of study, by Classification of Instructional Programs (CIP), is Business, management and public administration - Female | 25% | 2054/2030×100 | 1737/1713×100 | 2054/2030×100 |
| pct_cip_buisiness_f | Percentage of population aged 15+ whose major filed of study, by Classification of Instructional Programs (CIP), is Business, management and public administration - Male | 25% | 2054/2030×100 | 1737/1713×100 | 2054/2030×100 |
| pct_cip_physical_t | Percentage of population aged 15+ whose major filed of study, by Classification of Instructional Programs (CIP), is Physical and life sciences and technologies - Total | 25% | 2058/2030×100 | 1741/1713×100 | 2058/2030×100 |
| pct_cip_physical_f | Percentage of population aged 15+ whose major filed of study, by Classification of Instructional Programs (CIP), is Physical and life sciences and technologies - Female | 25% | 2058/2030×100 | 1741/1713×100 | 2058/2030×100 |
| pct_cip_physical_m | Percentage of population aged 15+ whose major filed of study, by Classification of Instructional Programs (CIP), is Physical and life sciences and technologies - Male | 25% | 2058/2030×100 | 1741/1713×100 | 2058/2030×100 |
| pct_cip_math_t | Percentage of population aged 15+ whose major filed of study, by Classification of Instructional Programs (CIP), is Mathematics, computer and information sciences - Total | 25% | 2064/2030×100 | 1747/1713×100 | 2064/2030×100 |
| pct_cip_math_f | Percentage of population aged 15+ whose major filed of study, by Classification of Instructional Programs (CIP), is Mathematics, computer and information sciences - Female | 25% | 2064/2030×100 | 1747/1713×100 | 2064/2030×100 |
| pct_cip_math_m | Percentage of population aged 15+ whose major filed of study, by Classification of Instructional Programs (CIP), is Mathematics, computer and information sciences - Male | 25% | 2064/2030×100 | 1747/1713×100 | 2064/2030×100 |

| | | | | | |
|---|---|---|---|---|---|
| pct_cip_architecture_t | Percentage of population aged 15+ whose major filed of study, by Classification of Instructional Programs (CIP), is Architecture, engineering, and related technologies - Total | 25% | 2069/2030×100 | 1752/1713×100 | 2069/2030×100 |
| pct_cip_architecture_f | The percentage of population aged 15+ whose major filed of study, by Classification of Instructional Programs (CIP), is Architecture, engineering, and related technologies - Female | 25% | 2069/2030×100 | 1752/1713×100 | 2069/2030×100 |
| pct_cip_architecture_m | Percentage of population aged 15+ whose major filed of study, by Classification of Instructional Programs (CIP), is Architecture, engineering, and related technologies - Male (25% sample data) | 25% | 2069/2030×100 | 1752/1713×100 | 2069/2030×100 |
| pct_cip_agriculture_t | Percentage of population aged 15+ whose major filed of study, by Classification of Instructional Programs (CIP), is Agriculture, natural resources and conservation - Total | 25% | 2077/2030×100 | 1760/1713×100 | 2077/2030×100 |
| pct_cip_agriculture_f | Percentage of population aged 15+ whose major filed of study, by Classification of Instructional Programs (CIP), is Agriculture, natural resources and conservation - Female | 25% | 2077/2030×100 | 1760/1713×100 | 2077/2030×100 |
| pct_cip_agriculture_m | Percentage of population aged 15+ whose major filed of study, by Classification of Instructional Programs (CIP), is Agriculture, natural resources and conservation - Male | 25% | 2077/2030×100 | 1760/1713×100 | 2077/2030×100 |
| pct_cip_health_t | Percentage of population aged 15+ whose major filed of study, by Classification of Instructional Programs (CIP), is Health and related fields - Total | 25% | 2080/2030×100 | 1763/1713×100 | 2080/2030×100 |
| pct_cip_health_f | Percentage of population aged 15+ whose major filed of study, by Classification of Instructional Programs (CIP), is Health and related fields - Female | 25% | 2080/2030×100 | 1763/1713×100 | 2080/2030×100 |
| pct_cip_health_m | Percentage of population aged 15+ whose major filed of study, by Classification of Instructional Programs (CIP), is Health and related fields - Male | 25% | 2080/2030×100 | 1763/1713×100 | 2080/2030×100 |
| pct_cip_personal_t | Percentage of population aged 15+ whose major filed of study, by Classification of Instructional Programs (CIP), is Personal, protective and transportation services - Total | 25% | 2086/2030×100 | 1767/1713×100 | 2086/2030×100 |

| | | | | | |
|---|---|---|---|---|---|
| pct_cip_personal_f | Percentage of population aged 15+ whose major filed of study, by Classification of Instructional Programs (CIP), is Personal, protective and transportation services - Female | 25% | 2086/2030×100 | 1767/1713×100 | 2086/2030×100 |
| pct_cip_personal_m | Percentage of population aged 15+ whose major filed of study, by Classification of Instructional Programs (CIP), is Personal, protective and transportation services - Male | 25% | 2086/2030×100 | 1767/1713×100 | 2086/2030×100 |
| pct_lf_participation_t | Labour force participation rate - Total | 25% | 2228 | 1870 | 2228 |
| pct_lf_participation_f | Labour force participation rate - Female | 25% | 2228 | 1870 | 2228 |
| pct_lf_participation_m | Labour force participation rate - Male | 25% | 2228 | 1870 | 2228 |
| pct_emp_t | Employment rate in population aged 15+ - Total | 25% | 2229 | 1871 | 2229 |
| pct_emp_f | Employment rate in population aged 15+ - Female | 25% | 2229 | 1871 | 2229 |
| pct_emp_m | Employment rate in population aged 15+ - Male | 25% | 2229 | 1871 | 2229 |
| pct_unemp_t | Unemployment rate in population aged 15+ - Total | 25% | 2230 | 1872 | 2230 |
| pct_unemp_f | Unemployment rate in population aged 15+ - Female | 25% | 2230 | 1872 | 2230 |
| pct_unemp_m | Unemployment rate in population aged 15+ - Male | 25% | 2230 | 1872 | 2230 |
| pct_self_emp_t | Percentage of labour force aged 15+ who are self-employed - Total | 25% | 2245/2237×100 | 1883/1879×100 | 2245/2237×100 |
| pct_self_emp_f | Percentage of labour force aged 15+ who are self-employed - Female | 25% | 2245/2237×100 | 1883/1879×100 | 2245/2237×100 |
| pct_self_emp_m | Percentage of labour force aged 15+ who are self-employed - Male | 25% | 2245/2237×100 | 1883/1879×100 | 2245/2237×100 |
| pct_noc_0_t | Percentage of labour force aged 15+ in Management occupations (NOC 0) - Total | 25% | 2249/2246×100 | 1887/1884×100 | 2249/2246×100 |
| pct_noc_0_f | Percentage of labour force aged 15+ in Management occupations (NOC 0) - Female | 25% | 2249/2246×100 | 1887/1884×100 | 2249/2246×100 |
| pct_noc_0_m | Percentage of labour force aged 15+ in Management occupations (NOC 0) - Male | 25% | 2249/2246×100 | 1887/1884×100 | 2249/2246×100 |
| pct_noc_1_t | Percentage of labour force aged 15+ in Business, finance and administration occupations occupations (NOC 1) - Total | 25% | 2250/2246×100 | 1888/1884×100 | 2250/2246×100 |
| pct_noc_1_f | Percentage of labour force aged 15+ in Business, finance and administration occupations occupations (NOC 1) - Female | 25% | 2250/2246×100 | 1888/1884×100 | 2250/2246×100 |

| | | | | | |
|---|---|---|---|---|---|
| pct_noc_1_m | Percentage of labour force aged 15+ in Business, finance and administration occupations occupations (NOC 1) - Male | 25% | 2250/2246×100 | 1888/1884×100 | 2250/2246×100 |
| pct_noc_2_t | Percentage of labour force aged 15+ in Natural and applied sciences and related occupations occupations (NOC 2) - Total | 25% | 2251/2246×100 | 1889/1884×100 | 2251/2246×100 |
| pct_noc_2_f | Percentage of labour force aged 15+ in Natural and applied sciences and related occupations occupations (NOC 2) - Female | 25% | 2251/2246×100 | 1889/1884×100 | 2251/2246×100 |
| pct_noc_2_m | Percentage of labour force aged 15+ in Natural and applied sciences and related occupations occupations (NOC 2) - Male | 25% | 2251/2246×100 | 1889/1884×100 | 2251/2246×100 |
| pct_noc_3_t | Percentage of labour force aged 15+ in health occupations (NOC 3) - Total | 25% | 2252/2246×100 | 1890/1884×100 | 2252/2246×100 |
| pct_noc_3_f | Percentage of labour force aged 15+ in health occupations (NOC 3) - Female | 25% | 2252/2246×100 | 1890/1884×100 | 2252/2246×100 |
| pct_noc_3_m | Percentage of labour force aged 15+ in health occupations (NOC 3) - Male | 25% | 2252/2246×100 | 1890/1884×100 | 2252/2246×100 |
| pct_noc_4_t | Percentage of labour force aged 15+ in education, law and social, community and government services occupations (NOC 4) - Total | 25% | 2253/2246×100 | 1891/1884×100 | 2253/2246×100 |
| pct_noc_4_f | The percentage of people aged 15+ in education, law and social, community and government services occupations (NOC 4) - Female | 25% | 2253/2246×100 | 1891/1884×100 | 2253/2246×100 |
| pct_noc_4_m | Percentage of labour force aged 15+ in education, law and social, community and government services occupations (NOC 4) - Male | 25% | 2253/2246×100 | 1891/1884×100 | 2253/2246×100 |
| pct_noc_5_t | Percentage of labour force aged 15+ in art, culture, recreation and sport occupations (NOC 5) - Total | 25% | 2254/2246×100 | 1892/1884×100 | 2254/2246×100 |
| pct_noc_5_f | Percentage of labour force aged 15+ in art, culture, recreation and sport occupations (NOC 5) - Female | 25% | 2254/2246×100 | 1892/1884×100 | 2254/2246×100 |
| pct_noc_5_m | Percentage of labour force aged 15+ in art, culture, recreation and sport occupations (NOC 5) - Male | 25% | 2254/2246×100 | 1892/1884×100 | 2254/2246×100 |
| pct_noc_6_t | Percentage of labour force aged 15+ in sales and service occupations (NOC 6) - Total | 25% | 2255/2246×100 | 1893/1884×100 | 2255/2246×100 |
| pct_noc_6_f | Percentage of labour force aged 15+ in sales and service occupations (NOC 6) - Female | 25% | 2255/2246×100 | 1893/1884×100 | 2255/2246×100 |
| pct_noc_6_m | Percentage of labour force aged 15+ in sales and service occupations (NOC 6) - Male | 25% | 2255/2246×100 | 1893/1884×100 | 2255/2246×100 |

| | | | | | |
|---|---|---|---|---|---|
| pct_noc_7_t | Percentage of labour force aged 15+ in trades, transport and equipment operators and related occupations (NOC 7) - Total | 25% | 2256/2246×100 | 1894/1884×100 | 2256/2246×100 |
| pct_noc_7_f | Percentage of labour force aged 15+ in trades, transport and equipment operators and related occupations (NOC 7) - Female | 25% | 2256/2246×100 | 1894/1884×100 | 2256/2246×100 |
| pct_noc_7_m | Percentage of labour force aged 15+ in trades, transport and equipment operators and related occupations (NOC 7) - Male | 25% | 2256/2246×100 | 1894/1884×100 | 2256/2246×100 |
| pct_noc_8_t | Percentage of labour force aged 15+ in natural resources, agriculture and related production occupations (NOC 8) - Total | 25% | 2257/2246×100 | 1895/1884×100 | 2257/2246×100 |
| pct_noc_8_f | Percentage of labour force aged 15+ in natural resources, agriculture and related production occupations (NOC 8) - Female | 25% | 2257/2246×100 | 1895/1884×100 | 2257/2246×100 |
| pct_noc_8_m | Percentage of labour force aged 15+ in natural resources, agriculture and related production occupations (NOC 8) - Male | 25% | 2257/2246×100 | 1895/1884×100 | 2257/2246×100 |
| pct_noc_9_t | Percentage of labour force aged 15+ in manufacturing and utilities occupations (NOC 9) - Total | 25% | 2258/2246×100 | 1896/1884×100 | 2258/2246×100 |
| pct_noc_9_f | Percentage of labour force aged 15+ in manufacturing and utilities occupations (NOC 9) - Female | 25% | 2258/2246×100 | 1896/1884×100 | 2258/2246×100 |
| pct_noc_9_m | Percentage of labour force aged 15+ in manufacturing and utilities occupations (NOC 9) - Male | 25% | 2258/2246×100 | 1896/1884×100 | 2258/2246×100 |
| pct_naics_11_t | Percentage of labour force aged 15+ in agriculture, forestry, fishing and hunting industry (NAICS 11) - Total | 25% | 2262/2259×100 | 1900/1897×100 | 2262/2259×100 |
| pct_naics_11_f | Percentage of labour force aged 15+ in agriculture, forestry, fishing and hunting industry (NAICS 11) - Female | 25% | 2262/2259×100 | 1900/1897×100 | 2262/2259×100 |
| pct_naics_11_m | Percentage of labour force aged 15+ in agriculture, forestry, fishing and hunting industry (NAICS 11) - Male | 25% | 2262/2259×100 | 1900/1897×100 | 2262/2259×100 |
| pct_naics_21_t | Percentage of labour force aged 15+ in mining, quarrying, and oil and gas extraction industry (NAICS 21) - Total | 25% | 2263/2259×100 | 1901/1897×100 | 2263/2259×100 |
| pct_naics_21_f | Percentage of labour force aged 15+ in mining, quarrying, and oil and gas extraction industry (NAICS 21) - Female | 25% | 2263/2259×100 | 1901/1897×100 | 2263/2259×100 |

| | | | | | |
|---|---|---|---|---|---|
| pct_naics_21_m | Percentage of labour force aged 15+ in mining, quarrying, and oil and gas extraction industry (NAICS 21) - Male | 25% | 2263/2259×100 | 1901/1897×100 | 2263/2259×100 |
| pct_naics_22_t | Percentage of labour force aged 15+ in utilities industry (NAICS 22) - Total | 25% | 2264/2259×100 | 1902/1897×100 | 2264/2259×100 |
| pct_naics_22_f | Percentage of labour force aged 15+ in utilities industry (NAICS 22) - Female | 25% | 2264/2259×100 | 1902/1897×100 | 2264/2259×100 |
| pct_naics_22_m | Percentage of labour force aged 15+ in utilities industry (NAICS 22) - Male | 25% | 2264/2259×100 | 1902/1897×100 | 2264/2259×100 |
| pct_naics_23_t | Percentage of labour force aged 15+ in construction industry (NAICS 23) - Total | 25% | 2265/2259×100 | 1903/1897×100 | 2265/2259×100 |
| pct_naics_23_f | Percentage of labour force aged 15+ in construction industry (NAICS 23) - Female | 25% | 2265/2259×100 | 1903/1897×100 | 2265/2259×100 |
| pct_naics_23_m | Percentage of labour force aged 15+ in construction industry (NAICS 23) - Male | 25% | 2265/2259×100 | 1903/1897×100 | 2265/2259×100 |
| pct_naics_31to33_t | Percentage of labour force aged 15+ in manufacturing industry (NAICS 31-33) - Total | 25% | 2266/2259×100 | 1904/1897×100 | 2266/2259×100 |
| pct_naics_31to33_f | Percentage of labour force aged 15+ in manufacturing industry (NAICS 31-33) - Female | 25% | 2266/2259×100 | 1904/1897×100 | 2266/2259×100 |
| pct_naics_31to33_m | Percentage of labour force aged 15+ in manufacturing industry (NAICS 31-33) - Male | 25% | 2266/2259×100 | 1904/1897×100 | 2266/2259×100 |
| pct_naics_41_t | Percentage of labour force aged 15+ in wholesale trade industry (NAICS 41) - Total | 25% | 2267/2259×100 | 1905/1897×100 | 2267/2259×100 |
| pct_naics_41_f | Percentage of labour force aged 15+ in wholesale trade industry (NAICS 41) - Female | 25% | 2267/2259×100 | 1905/1897×100 | 2267/2259×100 |
| pct_naics_41_m | Percentage of labour force aged 15+ in wholesale trade industry (NAICS 41) - Male | 25% | 2267/2259×100 | 1905/1897×100 | 2267/2259×100 |
| pct_naics_44to45_t | Percentage of labour force aged 15+ in retail trade industry (NAICS 44-45) - Total | 25% | 2268/2259×100 | 1906/1897×100 | 2268/2259×100 |
| pct_naics_44to45_f | Percentage of labour force aged 15+ in retail trade industry (NAICS 44-45) - Female | 25% | 2268/2259×100 | 1906/1897×100 | 2268/2259×100 |
| pct_naics_44to45_m | Percentage of labour force aged 15+ in retail trade industry (NAICS 44-45) - Male | 25% | 2268/2259×100 | 1906/1897×100 | 2268/2259×100 |
| pct_naics_48to49_t | Percentage of labour force aged 15+ in transportation and warehousing industry (NAICS 48-49) - Total | 25% | 2269/2259×100 | 1907/1897×100 | 2269/2259×100 |
| pct_naics_48to49_f | Percentage of labour force aged 15+ in transportation and warehousing industry (NAICS 48-49) - Female | 25% | 2269/2259×100 | 1907/1897×100 | 2269/2259×100 |

| | | | | | |
|---|---|---|---|---|---|
| pct_naics_48to49_m | Percentage of labour force aged 15+ in transportation and warehousing industry (NAICS 48-49) - Male | 25% | 2269/2259×100 | 1907/1897×100 | 2269/2259×100 |
| pct_naics_51_t | Percentage of labour force aged 15+ in information and cultural industries (NAICS 51) - Total | 25% | 2270/2259×100 | 1908/1897×100 | 2270/2259×100 |
| pct_naics_51_f | Percentage of labour force aged 15+ in information and cultural industries (NAICS 51) - Female | 25% | 2270/2259×100 | 1908/1897×100 | 2270/2259×100 |
| pct_naics_51_m | Percentage of labour force aged 15+ in information and cultural industries (NAICS 51) - Male | 25% | 2270/2259×100 | 1908/1897×100 | 2270/2259×100 |
| pct_naics_52_t | Percentage of labour force aged 15+ in finance and insurance industry (NAICS 52) - Total | 25% | 2271/2259×100 | 1909/1897×100 | 2271/2259×100 |
| pct_naics_52_f | Percentage of labour force aged 15+ in finance and insurance industry (NAICS 52) - Female | 25% | 2271/2259×100 | 1909/1897×100 | 2271/2259×100 |
| pct_naics_52_m | Percentage of labour force aged 15+ in finance and insurance industry (NAICS 52) - Male | 25% | 2271/2259×100 | 1909/1897×100 | 2271/2259×100 |
| pct_naics_53_t | Percentage of labour force aged 15+ in real estate and rental and leasing industry (NAICS 53) - Total | 25% | 2272/2259×100 | 1910/1897×100 | 2272/2259×100 |
| pct_naics_53_f | Percentage of labour force aged 15+ in real estate and rental and leasing industry (NAICS 53) - Female | 25% | 2272/2259×100 | 1910/1897×100 | 2272/2259×100 |
| pct_naics_53_m | Percentage of labour force aged 15+ in real estate and rental and leasing industry (NAICS 53) - Male | 25% | 2272/2259×100 | 1910/1897×100 | 2272/2259×100 |
| pct_naics_54_t | Percentage of labour force aged 15+ in professional, scientific and technical services industry (NAICS 54) - Total | 25% | 2273/2259×100 | 1911/1897×100 | 2273/2259×100 |
| pct_naics_54_f | Percentage of labour force aged 15+ in professional, scientific and technical services industry (NAICS 54) - Female | 25% | 2273/2259×100 | 1911/1897×100 | 2273/2259×100 |
| pct_naics_54_m | Percentage of labour force aged 15+ in professional, scientific and technical services industry (NAICS 54) - Male | 25% | 2273/2259×100 | 1911/1897×100 | 2273/2259×100 |
| pct_naics_55_t | Percentage of labour force aged 15+ in management of companies and enterprises industry (NAICS 55) - Total | 25% | 2274/2259×100 | 1912/1897×100 | 2274/2259×100 |
| pct_naics_55_f | Percentage of labour force aged 15+ in management of companies and enterprises industry (NAICS 55) - Female | 25% | 2274/2259×100 | 1912/1897×100 | 2274/2259×100 |
| pct_naics_55_m | Percentage of labour force aged 15+ in management of companies and enterprises industry (NAICS 55) - Male | 25% | 2274/2259×100 | 1912/1897×100 | 2274/2259×100 |

| | | | | | |
|---|---|---|---|---|---|
| pct_naics_56_t | Percentage of labour force aged 15+ in administrative and support, waste management and remediation services industry (NAICS 56) - Total | 25% | 2275/2259×100 | 1913/1897×100 | 2275/2259×100 |
| pct_naics_56_f | Percentage of labour force aged 15+ in administrative and support, waste management and remediation services industry (NAICS 56) - Female | 25% | 2275/2259×100 | 1913/1897×100 | 2275/2259×100 |
| pct_naics_56_m | Percentage of labour force aged 15+ in administrative and support, waste management and remediation services industry (NAICS 56) - Male | 25% | 2275/2259×100 | 1913/1897×100 | 2275/2259×100 |
| pct_naics_61_t | Percentage of labour force aged 15+ in educational services industry (NAICS 61) - Total | 25% | 2276/2259×100 | 1914/1897×100 | 2276/2259×100 |
| pct_naics_61_f | Percentage of labour force aged 15+ in educational services industry (NAICS 61) - Female | 25% | 2276/2259×100 | 1914/1897×100 | 2276/2259×100 |
| pct_naics_61_m | Percentage of labour force aged 15+ in educational services industry (NAICS 61) - Male | 25% | 2276/2259×100 | 1914/1897×100 | 2276/2259×100 |
| pct_naics_62_t | Percentage of labour force aged 15+ in health care and social assistance industry (NAICS 62) - Total | 25% | 2277/2259×100 | 1915/1897×100 | 2277/2259×100 |
| pct_naics_62_f | Percentage of labour force aged 15+ in health care and social assistance industry (NAICS 62) - Female | 25% | 2277/2259×100 | 1915/1897×100 | 2277/2259×100 |
| pct_naics_62_m | Percentage of labour force aged 15+ in health care and social assistance industry (NAICS 62) - Male | 25% | 2277/2259×100 | 1915/1897×100 | 2277/2259×100 |
| pct_naics_71_t | Percentage of labour force aged 15+ in arts, entertainment and recreation industry (NAICS 71) - Total | 25% | 2278/2259×100 | 1916/1897×100 | 2278/2259×100 |
| pct_naics_71_f | Percentage of labour force aged 15+ in arts, entertainment and recreation industry (NAICS 71) - Female | 25% | 2278/2259×100 | 1916/1897×100 | 2278/2259×100 |
| pct_naics_71_m | Percentage of labour force aged 15+ in arts, entertainment and recreation industry (NAICS 71) - Male | 25% | 2278/2259×100 | 1916/1897×100 | 2278/2259×100 |
| pct_naics_72_t | Percentage of labour force aged 15+ in accommodation and food services industry (NAICS 72) - Total | 25% | 2279/2259×100 | 1917/1897×100 | 2279/2259×100 |
| pct_naics_72_f | Percentage of labour force aged 15+ in accommodation and food services industry (NAICS 72) - Female | 25% | 2279/2259×100 | 1917/1897×100 | 2279/2259×100 |
| pct_naics_72_m | Percentage of labour force aged 15+ in accommodation and food services industry (NAICS 72) - Male | 25% | 2279/2259×100 | 1917/1897×100 | 2279/2259×100 |

| | | | | | |
|---|---|---|---|---|---|
| pct_naics_81_t | Percentage of labour force aged 15+ in other services (except public administration) industry (NAICS 81) - Total | 25% | 2280/2259×100 | 1918/1897×100 | 2280/2259×100 |
| pct_naics_81_f | Percentage of labour force aged 15+ in other services (except public administration) industry (NAICS 81) - Female | 25% | 2280/2259×100 | 1918/1897×100 | 2280/2259×100 |
| pct_naics_81_m | Percentage of labour force aged 15+ in other services (except public administration) industry (NAICS 81) - Male | 25% | 2280/2259×100 | 1918/1897×100 | 2280/2259×100 |
| pct_naics_91_t | Percentage of labour force aged 15+ in public administration industry (NAICS 91) - Total | 25% | 2281/2259×100 | 1919/1897×100 | 2281/2259×100 |
| pct_naics_91_f | Percentage of labour force aged 15+ in public administration industry (NAICS 91) - Female | 25% | 2281/2259×100 | 1919/1897×100 | 2281/2259×100 |
| pct_naics_91_m | Percentage of labour force aged 15+ in public administration industry (NAICS 91) - Male | 25% | 2281/2259×100 | 1919/1897×100 | 2281/2259×100 |

Notes: [1]The sample from which the variable is derived. The sample can be 100% (from long-form census) or 25% (from short-form census). [2]We have assigned the Profile IDs for year 2011 as Census Profile 2011 and NHS Profile only provides the charactheristics, without an accompanied uniqe id for them. [3] The 2016 Profile ID is the *Member ID: Profile of Dissemination Areas (2247)* variable in Census Profile 2016. [4]The 2021 Profile ID is the *CHARACTERISTIC_ID* in Census Profile 2021. [5]`year' can take 11, 16, and 21, which indicates variables corresponding to 2011, 2016, and 2021 boundaries, respectively. [6]NA: Not Applicable.
